# Tau isoform imbalance and aggregation are pathological hallmarks of X-linked dystonia-parkinsonism

**DOI:** 10.64898/2026.07.23.26358614

**Authors:** Charles Jourdan Reyes, Aloysius Domingo, Ellen B. Penney, Ean Norenberg, Justin Han, Micaela G. Murcar, Christine A. Vaine, Nicolas A. Bravo-Vasquez, Hoang-Dai Tran, Noé Quittot, Anastasie Mate de Gerando, Nil Saez-Calveras, Yogesh Tak, Rachita Yadav, Dadi Gao, Siddarth Reed, Serkan Erdin, Nandini Ramesh, Benjamin Wymann, Aaron Held, Ranee Zara Monsanto, Laura Moran, Hayden Wheeler, Yin Yin Ruan, Grant Griesman, Grace Anne Field, Chao-Zong Lee, Gelbert Crescencio, Matthew Nolan, John Lemanski, Kathryn O’Keefe, Bimal Jana, Cara Fernandez-Cerado, M. Salvie Velasco-Andrada, Gierold Paul A. Legarda, Niecy Grace Ganza-Bautista, Michelle Sy, Madison Hincher, Tiziana Petrozziello, Pia Kivisäkk, Ghazaleh Sadri-Vakili, Edwin L. Muñoz, Mark Angelo C. Ang, Cid Czarina E. Diesta, Criscely Go, Mark W. Albers, Steven Arnold, Brian J. Wainger, Rachel E. Bennett, Marc I. Diamond, Jeffrey W. Miller, Bradley T. Hyman, Nutan Sharma, Laurie J. Ozelius, Michael E. Talkowski, D. Cristopher Bragg, Clotilde Lagier-Tourenne

**Author notes:** Correspondence (M.E.T), (D.C.B.), (C.L.-T.). Authors contributed equally (Charles Jourdan Reyes, Aloysius Domingo, Ellen B. Penney).

## Abstract

Tauopathies encompass diverse neurodegenerative diseases unified by aberrant patterns of tau deposition in brain. Although most appear sporadic, some are linked to genetic etiologies that offer unique mechanistic insights. Here we report that X-linked Dystonia-Parkinsonism (XDP), caused by a non-coding retrotransposon-associated repeat insertion in *TAF1*, involves a significant imbalance of tau isoforms and the accumulation of hyperphosphorylated, four-repeat tau in the brain. In striatal tissue, both misfolded tau accumulation, predominantly in astrocytes, and *MAPT* exon 10 inclusion correlated with repeat length within the causal insertion. Transcriptomic profiling across brain regions revealed dysregulation of known tau-related pathways. Levels of phosphorylated tau181, glial fibrillary acidic protein, and neurofilament light chain were elevated in patient plasma and discriminated XDP from controls. These findings implicate defective tau proteostasis as a key pathogenic mechanism and position XDP as a genetic model for uncovering cellular drivers that may disrupt tau in other more common neurodegenerative diseases.

## INTRODUCTION

Human tauopathies are a broad group of neurodegenerative diseases characterized by the aggregation and propagation of the microtubule-associated protein, tau^1^. Under physiological conditions, tau is predominantly localized to neuronal axons, but in the pathological state, it is redistributed to cell bodies, where it becomes increasingly hyperphosphorylated to form insoluble deposits^2^. Across diverse tauopathies, patterns of tau phosphorylation, aggregation, and seeding correspond to the severity of neurodegeneration and disease progression^3–6^, and despite their clinical heterogeneity, many aspects of the observed pathologies are common features among these neurodegenerative conditions. These similarities raise the possibility that convergent molecular mechanisms may underlie tau dysregulation across distinct disorders and that elucidating such pathways may reveal therapeutic opportunities that are widely applicable to tau-related syndromes. Toward that end, identifying disorders with known genetic etiologies that involve disturbances in tau can offer a powerful approach, because the genetically defined patient cohorts and corresponding experimental models can enable dissection of cellular mechanisms that may be generalizable to more common, sporadic forms of disease^7–13^.

In this study, we investigated neuropathological features associated with X-linked Dystonia-Parkinsonism (XDP), a rare Mendelian neurodegenerative disease, and identified region-specific disruptions in tau homeostasis, including aberrant tau accumulation and altered phosphorylation patterns. XDP is endemic to Panay Island in the Philippines due to a genetic founder effect^14^, and affected individuals develop a severe adult-onset movement disorder consisting of both hyper- and hypokinetic features^15–17^. The disease is caused by the insertion of a SINE-VNTR-*Alu* (SVA)-type retrotransposon within intron 32 of *TAF1*^18^, which encodes the largest subunit of the TFIID complex required for RNA polymerase II-mediated transcription. The SVA contains a polymorphic (CCCTCT)_n_ repeat motif, the length of which is inversely correlated to age at disease onset^19,20^. The inserted retroelement induces multiple disruptions at the surrounding locus, including increased retention of *TAF1* intron 32 and aberrant alternative splicing^18^, formation of G-quadruplexes^21^ and R-loops^22^, as well as epigenetic remodeling that establishes altered heterochromatin domains^23,24^. Together, these effects are thought to disrupt local transcription of *TAF1* and reduce the full-length mRNA^18^.

The mechanisms underlying neurodegeneration in XDP remain unclear, and to date there have been only limited neuropathological studies examining post-mortem brain tissue. Given that protein aggregation and impaired proteostasis are common pathogenic features across many neurodegenerative disorders, we hypothesized that neuronal loss in XDP may be similarly associated with aberrant protein accumulation. Consistent with that hypothesis, we previously performed transcriptomic profiling of XDP patient-derived cellular models that revealed dysregulation of gene sets linked to protein quality control pathways, including the autophagy-lysosome and ubiquitin-proteosome pathways^25^, as well as activation of the unfolded protein response^18^. While prior neuropathological studies of post-mortem cases have primarily focused on medium spiny neurons loss in the striatum reminiscent of the lesion in Huntington’s disease (HD)^26,27^, neuroimaging studies across different stages of XDP progression have detected alterations in additional brain regions. Specifically, the observed patterns indicate abnormalities in anatomically connected regions, including thinning of the frontal and temporal cortices, grey matter pathology in the cerebellum, and volume loss in the substantia nigra^28,29^, whereas the occipital and parietal cortices, which lack direct striatal projections, appear relatively preserved^30^. This network-selective pattern of degeneration suggests that XDP may involve the propagation of a pathogenic substrate through anatomically connected circuits, consistent with the prion-like spreading of misfolded proteins^31^. Collectively, these observations support a model in which impaired proteostasis in XDP leads to a neurodegenerative process encompassing more brain regions than previously recognized.

To determine whether XDP exhibits molecular and pathological signatures of a proteinopathy, we analyzed an extensive collection of XDP and control post-mortem brains and profiled plasma from a large cohort of patients and unaffected individuals. Integrative neuropathological and transcriptomic analyses of brain tissue revealed pathological inclusions composed of hyperphosphorylated, predominantly astrocytic four-repeat (4R) tau isoforms. In plasma from patients with XDP, levels of phosphorylated tau (p-tau181) and glial fibrillary acidic protein (GFAP), a marker of reactive astrogliosis, were significantly elevated and effectively discriminated XDP from control samples. Collectively, these findings identify a previously unrecognized role for tau splicing imbalance and aggregation as potential drivers of neuronal injury in XDP and indicate a link between the genetic lesion in *TAF1* and tau proteostasis. These results broaden the conceptual framework of XDP, suggesting that tau-lowering or proteostasis-restoring therapies may offer new avenues for treatment in this disorder while establishing it as a genetic model to uncover pathways leading to tau-mediated toxicity.

## RESULTS

### XDP shows brain region-specific disturbances in tau proteostasis

To systematically describe pathological and transcriptomic alterations in XDP brains, we accessed a large collection of post-mortem specimens from patients with XDP (n=42) and controls (n=16) from the Philippines (**Figure 1A**)^32^. Pertinent clinical, demographic, and genetic information for all donors is summarized in **Table S1**. In this cohort, the XDP-specific hexameric (CCCTCT)_n_ repeat length ranged from 34 to 52, and age at disease onset ranged from 26 to 58 years. Consistent with previous findings^33^, (CCCTCT)_n_ repeat length was inversely correlated with age at disease onset (r=-0.78, *p*=4.0e-09), and age at death (r=-0.79, *p*=6.0e-10), but not disease duration (r=-0.06, *p*=0.70) (**Figure S1**). Males with XDP were younger at death relative to Filipino controls (*p*=0.04). Post-mortem intervals and other technical factors (i.e., RNA integrity) were not significantly different between cohorts (**Table S1**).

**Figure 1.**
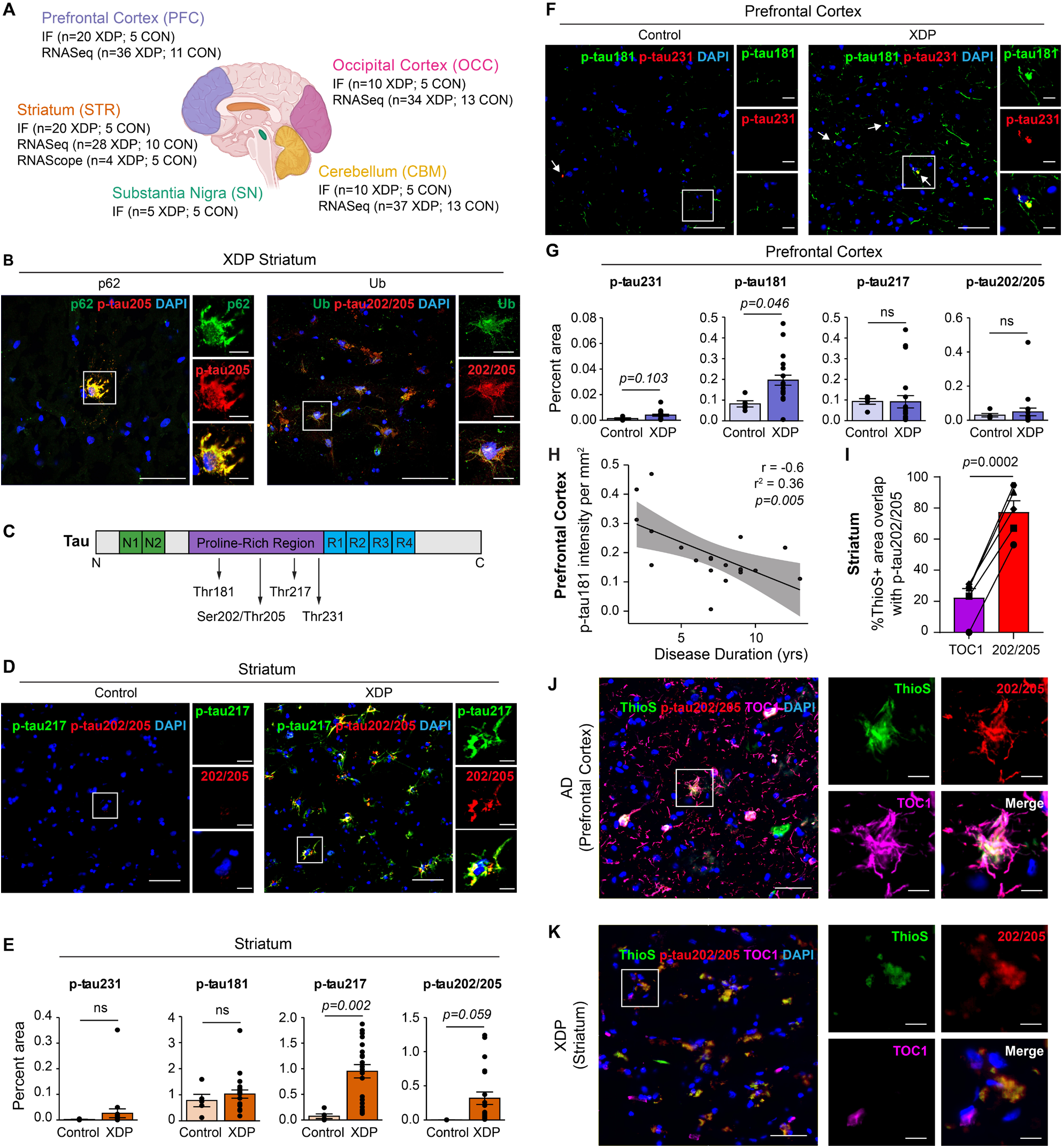
Brain region-specific accumulation of phospho-tau pathology in XDP brains. **A**, Schematic diagram of brain regions analyzed, including the prefrontal cortex (PFC), occipital cortex (OCC), striatum (STR), substantia nigra (SN), and cerebellum (CBM). Sample sizes for immunofluorescence (IF), RNA sequencing (RNA-seq), and RNAscope fluorescence *in situ* hybridization are indicated for each region. **B**, Representative confocal images of XDP striatum stained for p62 or ubiquitin (Ub) together with p-tau202/205 (AT8). Insets show higher-magnification views of boxed regions, demonstrating co-localization of phospho-tau with p62 and ubiquitin. **C**, Schematic diagram of tau protein domains and major phosphorylation sites analyzed, including p-tau181, p-tau217, p-tau202/205 (AT8), and p-tau231. **D**, Representative confocal images of control and XDP striatum stained for p-tau217 and p-tau202/205 (AT8). Insets show higher-magnification views of boxed regions. **E**, Quantification of phospho-tau burden in striatum shows increased p-tau217 (*p*=0.002) and p-tau202/205 (*p*=0.059) in XDP, with no significant differences in p-tau181 or p-tau231. **F**, Representative images of control and XDP prefrontal cortex stained for p-tau181 and p-tau231. Insets show higher-magnification views. **G**, Quantification of phospho-tau burden in prefrontal cortex shows increased p-tau181 (*p*=0.046) in XDP, with no significant changes in p-tau231, p-tau217, or p-tau202/205. **H**, p-tau181 immunoreactivity in prefrontal cortex inversely correlates with disease duration (r=−0.6, r^2^=0.36, *p*=0.005). **I**, Quantification of Thioflavin S (ThioS)-positive area overlapping with TOC1 or p-tau202/205 (AT8) in striatum shows greater overlap with p-tau202/205 compared to TOC1 (*p*=0.0002), indicating limited fibrillar tau formation. **J-K**, Representative confocal images of AD prefrontal cortex and XDP striatum stained for ThioS, p-tau202/205 (AT8), and TOC1. AD prefrontal cortex shows abundant ThioS-positive fibrillar aggregates co-localizing with p-tau202/205, whereas XDP striatum shows sparse ThioS labeling with predominant TOC1-positive, non-fibrillar oligomeric tau species. Insets show higher-magnification views. Data are mean ± s.e.m.; each point represents one case. Statistical significance is indicated in panels. Scale bars, 50 μm (overview images) and 10 μm (insets).

We first examined whether XDP brains showed evidence of impaired protein clearance or generalized protein aggregation by assessing the distribution of ubiquitin and p62, canonical markers of impaired protein degradation through the ubiquitin–proteasome and autophagy–lysosome systems, respectively. Immunofluorescence staining in XDP (n=20) and control brains (n=5) identified an increase in the total number of ubiquitinated aggregates in XDP striatum (*p*=0.03) (**Figure S2A, C**), an increase in p62-positive aggregates in the prefrontal cortex (*p*=0.03) (**Figure S2B, D**), and an increase in both types of aggregates in the cerebellum (*p*=0.01 [ubiquitin], *p*=1.2e-04 [p62]) (**Figure S2A-B, E, Table S2**). No differences in ubiquitin and p62 staining were observed in the substantia nigra or occipital cortex (**Figure S2A-B,F-G, Table S2**).

Based on these observations, we asked if the impaired proteostasis phenotype observed in XDP brains may be recapitulated in iPSC-differentiated neural stem cells (NSCs) derived from three sources: (i) patients with XDP (n=6); (ii) unaffected controls (n=5); and (iii) isogenic CRISPR/Cas9-edited XDP lines in which the SVA insertion in *TAF1* was excised (ΔSVA; n=6) and XDP-associated alternative splicing signatures ameliorated^18^. Cells were treated with increasing chloroquine concentrations, which inhibit autophagy by preventing autophagosome-lysosome fusion. Whereas p62 was not detected at baseline, all three genotypes exhibited dose-dependent accumulations in p62, with XDP cell lines showing significantly higher levels than both the unaffected control cells and ΔSVA lines, which were indistinguishable from each other (**Figure S2H-I**). Analysis of the dose-response curves by two-way ANOVA revealed significant effects of genotype [*p*<1.0e-04, F (2,552)] and treatment [(*p*<1.0e-04), F (3,552)]. These findings provide evidence of disrupted protein homeostasis in XDP brains and neural cell models.

To identify the core protein component of the inclusions observed in XDP brains, we performed double-label immunostaining using antibodies against proteins known to accumulate in neurodegenerative diseases. The ubiquitinated and p62-positive inclusions in XDP striatum were immunoreactive for hyperphosphorylated tau (**Figure 1B**), whereas no immunoreactivity was detected for pathological α-synuclein, TDP-43, or amyloid-β (**Figure S3A-I**).

Having identified tau as a component of protein inclusions in XDP, we next examined its phosphorylation state to determine if the pattern of tau modification within the parenchyma may suggest a progressive aggregation pattern related to disease stage. Tau is phosphorylated at multiple residues including Thr181 (p-tau181), Ser202 and Thr205 (p-tau202/205 [AT8]), Thr217 (p-tau217), and Thr231 (p-tau231) that accumulate at distinct stages of aggregation^34,35^, and patterns of tau phosphorylation correspond to the severity of neurodegeneration and disease progression in Alzheimer’s Disease (AD)^3,36^ (**Figure 1C**). We thus quantified immunoreactivity for each of these species in different brain regions and compared differences in phosphorylated tau after adjusting for age at death and within-individual variability. These analyses revealed increased levels of p-tau217 (*p*=2.1e-03) and p-tau202/205 (*p*=0.05) in the XDP striatum compared to controls (**Figure 1D-E, Table S2**). No accompanying changes in p-tau181 and p-tau231 were observed in this brain region (**Figures 1E and S4A**). In contrast, p-tau181 (*p*=0.04) levels were elevated in the XDP prefrontal cortex without changes in p-tau217 and p-tau202/205 (AT8) levels (**Figures 1F-G and S4B, Table S2**). The increase in p-tau181, consistent with its association with synaptic and neuritic tau in both normal aging and disease brains^3,37,38^, showed a significant inverse correlation with disease duration (r=-0.60, *p*=5.0e-03) (**Figure 1H**). Phospho-tau levels were unaltered in the substantia nigra and occipital cortex (**Figure S5A-F, Table S2**). Notably, the p-tau202/205 immunoreactive staining observed in XDP striatum colocalized with Thioflavin S (ThioS)-positive inclusions, indicating the presence of insoluble, fibrillar tau aggregates in this brain region (**Figure 1I-K**).

While the p62- and ubiquitin-positive inclusions in XDP striatum and prefrontal cortex appeared morphologically similar and contained hyperphosphorylated tau species, the inclusions detected within the cerebellum had a distinct pattern (**Figure S2E**). In these aggregates, ubiquitin and p62 were observed in concentric rings reminiscent of corpora amylacea, also known as “wasteosomes”, which are spherical, carbohydrate-rich structures that sequester misfolded proteins, damaged organelles, and other cellular debris^39^. To confirm this identity, we performed Periodic Acid-Schiff (PAS) staining, which labels the characteristic polysaccharide core, and detected abundant PAS-positive structures in the molecular layer of the XDP cerebellum (**Figure S6A**). Prior studies have shown that corpora amylacea contain early-stage tau species with molecular features distinct from end-stage aggregates, including immunoreactivity for total and hypo-phosphorylated tau but not for hyperphosphorylated p-tau202/205 species^40,41^. Consistent with these reports, double-label immunofluorescence for ubiquitin and total tau (based on Tau5 antibody labeling) demonstrated an accumulation of tau within these corpora amylacea (**Figure S6B**), and quantitative analysis of staining intensities showed that total tau levels were significantly increased in the molecular layer of the XDP cerebellum relative to corresponding control tissue (*p*=0.01) (**Figure S6C and 3A, Table S2**). However, this increase did not coincide with any evidence of hyperphosphorylated tau accumulation, consistent with previous descriptions of corpora amylacea (**Figure S6D**).

Together, these findings demonstrate region-specific disturbances in tau phosphorylation and proteostasis in XDP brains. We detected distinct phosphorylated tau species in the striatum and prefrontal cortex. In contrast, elevated tau in the cerebellum was localized within p62- and ubiquitin-positive corpora amylacea and did not appear to be hyperphosphorylated.

### Soluble misfolded tau species accumulate in striatal astrocytes and correlate with (CCCTCT)_n_ repeat length

Because soluble misfolded tau species have been implicated as key instigators of neurodegeneration in tauopathies^42–44^, we next probed for alterations in these specific forms in XDP and control brains. Tissue sections from both groups were immunostained with an antibody against total tau (Tau5) and a conformation-specific antibody that recognizes misfolded, oligomeric tau species (T22), previously validated to selectively detect tau oligomers independent of fibrillar aggregates^45–49^. The burden of soluble misfolded tau was significantly increased in the striatum of patients with XDP (p=5.9e-04), whereas the total tau levels in this region were comparable between groups (**Figure 2A-C, Table S2**). T22 staining was occasionally observed surrounding corpora amylacea in the cerebellum (**Figure S6C**), however, there was no significant increase of soluble misfolded tau in this region (**Figure 2B, Table S2**). Similarly, no significant differences in total or misfolded tau were detected in prefrontal cortex, substantia nigra, or occipital cortex (**Figures 2A-B, S7A-C, Table S2**).

**Figure 2.**
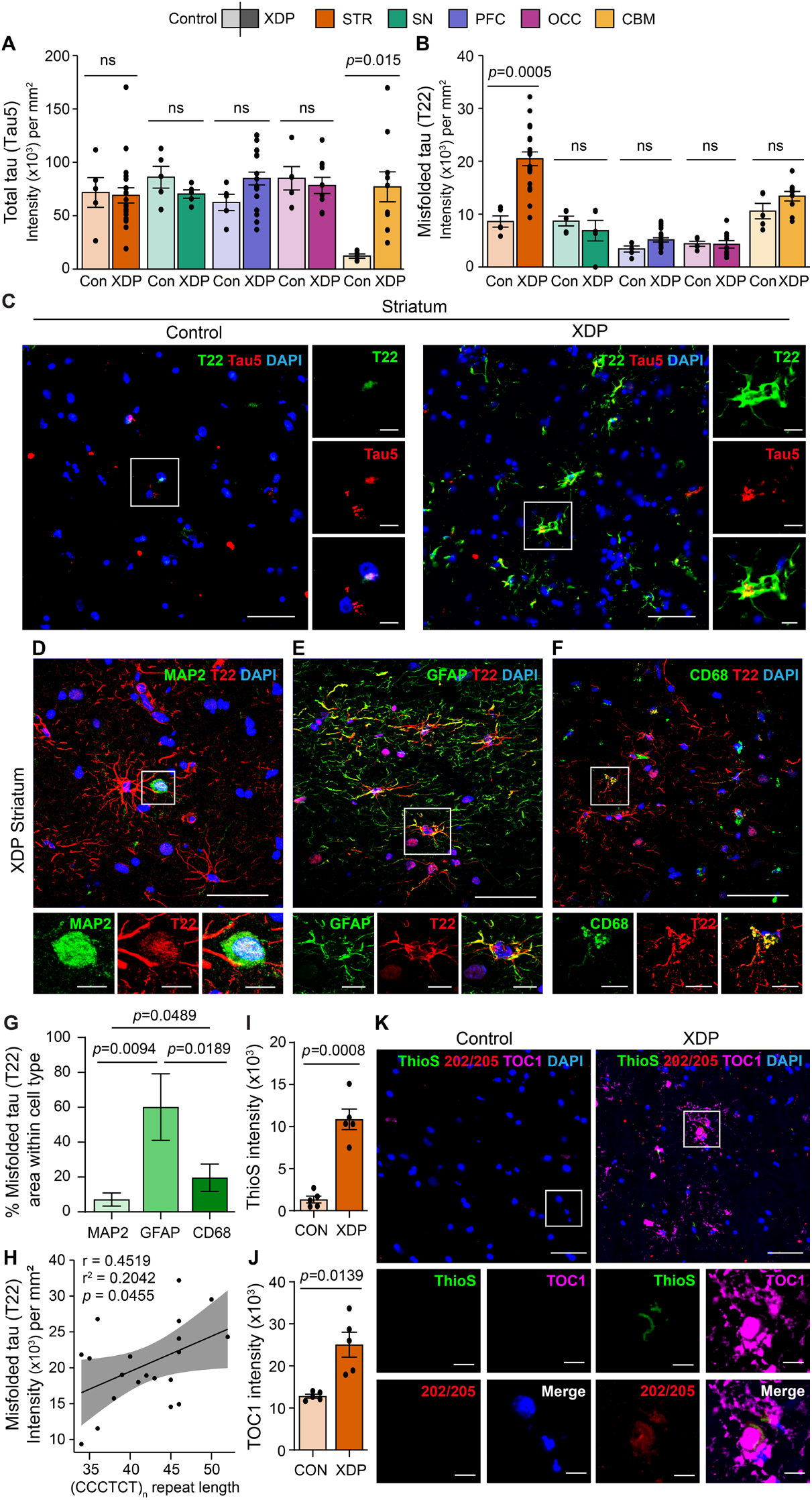
Soluble misfolded tau conformers are elevated in the XDP striatum, accumulate predominantly in astrocytes, and correlate with (CCCTCT)_n_ repeat length. **A-B**, Quantification of fluorescence intensity per mm^2^ shows that total tau (Tau5) levels remain unchanged, whereas misfolded oligomeric tau (T22) is significantly increased in XDP striatum (STR). **C**, Representative confocal images of control (n=5) and XDP (n=20) striatum stained for Tau5 and T22. **D-F**, Representative XDP striatum sections (n=5) co-stained for T22 with neuronal (MAP2) (**D**), astrocytic (GFAP) (**E**), and microglial (CD68) (**F**) markers, showing prominent astrocytic accumulation of soluble misfolded tau. **G**, Quantification of the fraction of T22-immunoreactive area within each cell type reveals preferential localization of misfolded tau to astrocytes compared to neurons and microglia. **H**, Pearson correlation analysis shows that T22 intensity per mm^2^ increases with (CCCTCT)_n_ repeat length within the TAF1 SVA insertion, but not with disease duration. **I-K**, Quantification of Thioflavin S (ThioS) and TOC1 immunofluorescence intensity shows increased TOC1-immunoreactive tau with limited ThioS labeling in XDP striatum, consistent with soluble, non-fibrillar, oligomeric tau species. Control striatum shows minimal staining with either marker. Data are mean ± s.e.m.; each point represents one case. Statistical significance is indicated in panels. Scale bars, 50 μm (overview images) and 10 μm (insets).

Having detected increased soluble misfolded tau in the XDP striatum, we next assessed its cellular distribution and relationship to the causal genetic variant in XDP. Double-label immunofluorescence for soluble misfolded tau (T22) combined with markers of neurons (MAP2), astrocytes (GFAP), and activated microglia (CD68) (n=5) revealed increased levels of misfolded tau in all three cell types (**Figure 2D-F**), with significant enrichment in astrocytes relative to neurons (p=0.01) and microglia (p=0.02) (**Figure 2G**), mirroring prior findings linking soluble misfolded tau to reactive astrocytosis and neuroinflammation^50–53^. Moreover, the levels of soluble misfolded tau in XDP striatum significantly correlated with the length of the pathogenic (CCCTCT)_n_ repeat sequence (r=0.45, p=0.04) (**Figure 2H**) with no residual correlations observed with age at disease onset, age at death, or disease duration in our models (data not shown). To validate the T22 staining results, we employed an independent tau oligomer-specific antibody (TOC1)^54^ in combination with ThioS staining and observed minimal overlap between TOC1- and ThioS-positive inclusions, consistent with the presence of soluble, higher-order tau assemblies rather than insoluble fibrillar aggregates (**Figure 2I-K**).

To assess whether the soluble misfolded tau species in XDP brain possess proteopathic seeding activity, we tested total brain lysates and insoluble protein fractions from the striatum and prefrontal cortex (n=5 XDP, 5 controls) using HEK293T v2L P301S tau biosensor cells^55^. Lysates from the prefrontal cortex of a patient with AD served as positive controls and produced robust FRET-positive signals indicative of strong seeding activity. In contrast, despite variability in seeding activity among individual cases, XDP total brain lysates and insoluble fractions did not elicit detectable seeding above control levels (**Figure 3A-B, S8**). This absence of measurable activity may reflect limited assay sensitivity to certain 4R tauopathy strains or conformational variants, given that the P301S biosensor lines preferentially responds to AD-type tau seeds^56^. Consequently, we next evaluated PBS-soluble striatal and prefrontal cortex lysates (n=5 XDP, 5 controls per brain region) using additional tau biosensor lines expressing wild-type 3R/4R and 4R/4R tau repeat domain constructs^57^, with two AD cases included as positive controls. Consistent with the P301S assay, there was no overall increase in tau seeding activity in XDP compared with controls (**Figure 3C-D**). These findings indicate that seed-competent tau species are not consistently enriched in XDP brain lysates. We also did not observe immunoreactivity in XDP striatum with the AD-tau-conformation-selective antibody GT-3858 (**Figure 3E**).

**Figure 3.**
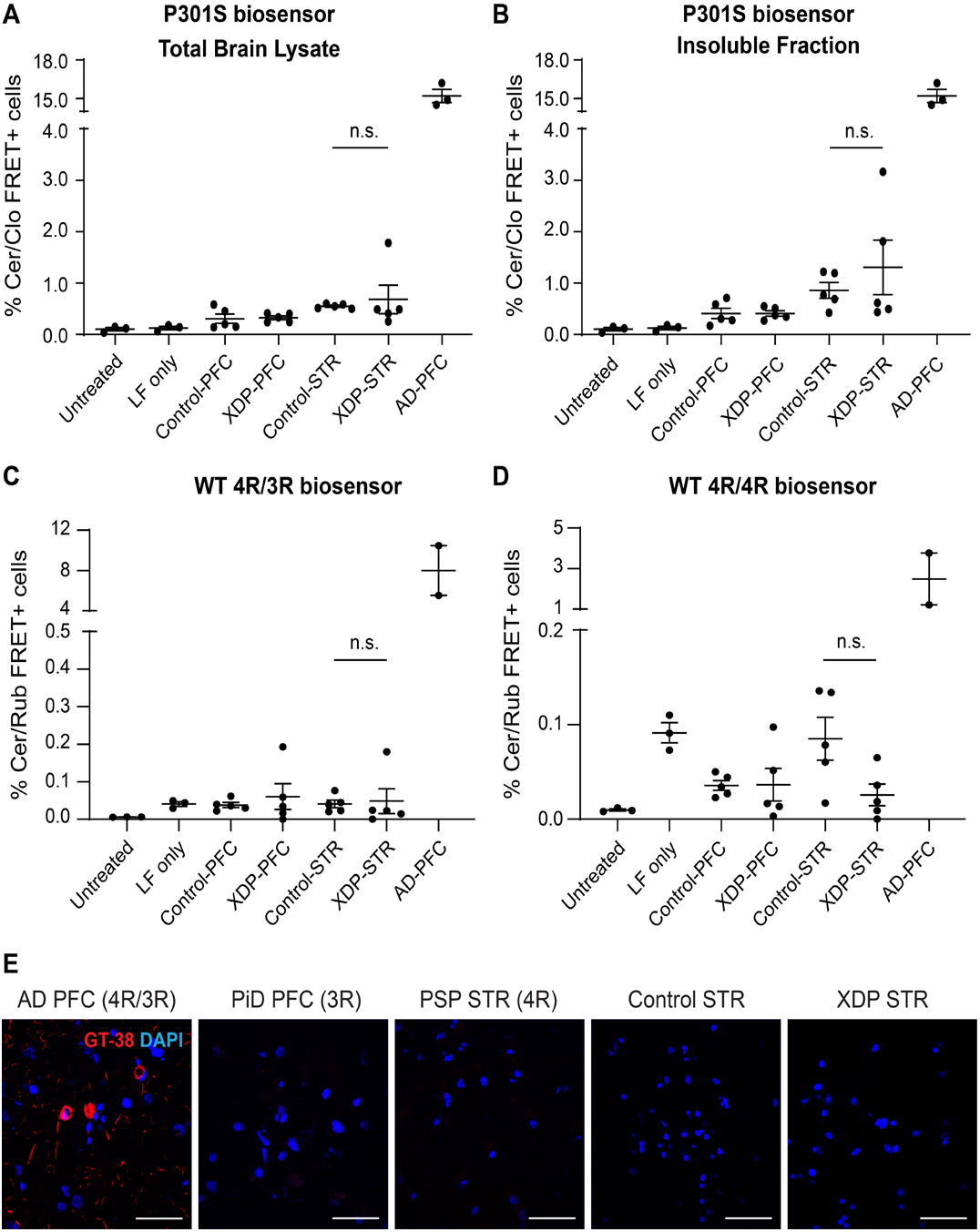
Absence of robust seed-competent and Alzheimer’s disease-type tau species in XDP brains. **A-D**, Tau seeding activity assessed using HEK293T tau biosensor cell lines. **A**, Total brain lysates and **B**, insoluble fractions prepared from the striatum (STR) and prefrontal cortex (PFC) of control (n=5) and X-linked dystonia-parkinsonism (XDP; n=5) brains were transfected into the commercial P301S tau biosensor cells. **C-D**, PBS-soluble lysates were additionally evaluated using wild-type 4R/3R (**C**) and wild-type 4R/4R (**D**) tau biosensor cell lines. Lipofectamine-only (LF only) served as the negative transfection control. Prefrontal cortex lysates from an Alzheimer’s disease (AD) case served as positive controls and exhibited robust tau seeding activity, whereas no overall increase in seeding activity was detected in XDP lysates across biosensor platforms. Seeding activity was quantified 48-72 h after transfection. **E**, Representative immunofluorescence images stained with GT-38, an antibody selective for the Alzheimer’s disease tau conformation, demonstrate robust immunoreactivity in AD prefrontal cortex (PFC) but no detectable labeling in XDP or control striatum (STR). Progressive supranuclear palsy (PSP) striatum (4R tauopathy) and Pick’s disease (PiD) prefrontal cortex (3R tauopathy) are shown as negative controls for GT-38 immunoreactivity. Data are presented as mean ± s.e.m.; each point represents one individual case. Statistical significance was assessed by ordinary two-way ANOVA. Scale bars, 50 μm.

Collectively, these experiments demonstrate that soluble misfolded tau accumulated preferentially in astrocytes within the XDP striatum, and its abundance correlated directly with genetic variation in the SVA retrotransposon sequence that is the cause of the disease. However, the tau species in XDP brain tissues generally lacked robust seeding activity in established biosensor assays.

### XDP brains display transcriptional signatures of neurodegeneration, tau aggregation, and neuroinflammation

To explore the global transcriptional consequences of increased tau burden in XDP brains, we performed RNA sequencing (RNA-seq) across multiple brain regions. RNA profiles from 41 XDP and 13 control donors were analyzed, with up to four regions sampled per individual (striatum, prefrontal cortex, cerebellum, occipital cortex; total libraries = 182, **Table S1**). The largest set of differentially expressed genes (DEGs) was identified in the striatum, which also exhibited the most prominent tau accumulation by immunostaining, with 1295 DEGs at false-discovery rate (FDR)-adjusted p<0.01 and |log2(fold-change)| >0.58 (**Figure 4A, Table S3**). Transcriptional changes were relatively more subtle in the cerebellum and prefrontal cortex at these same thresholds (222 and 187 DEGs, respectively), yet the DEGs observed in these tissues displayed a strong directional overlap with striatal DEGs that suggested shared transcriptional perturbations across tau-affected brain regions (largest direction-specific DEG pairwise enrichment p=1.4e-09) (**Table S3**). In contrast, the occipital cortex showed a remarkably small number of expression changes with only 7 DEGs at these same thresholds, consistent with the lack of detectable pathology in this brain region. To assess the robustness of the transcriptional changes predicted from these analyses, we compared these DEG analyses to bootstrapping of 50,000 iterations of the dataset. The distribution of DEGs in the case-control analyses demonstrated good calibration from the bootstrap analyses at these FDR and fold-change thresholds across all regions (**Figure S9A-C**).

**Figure 4.**
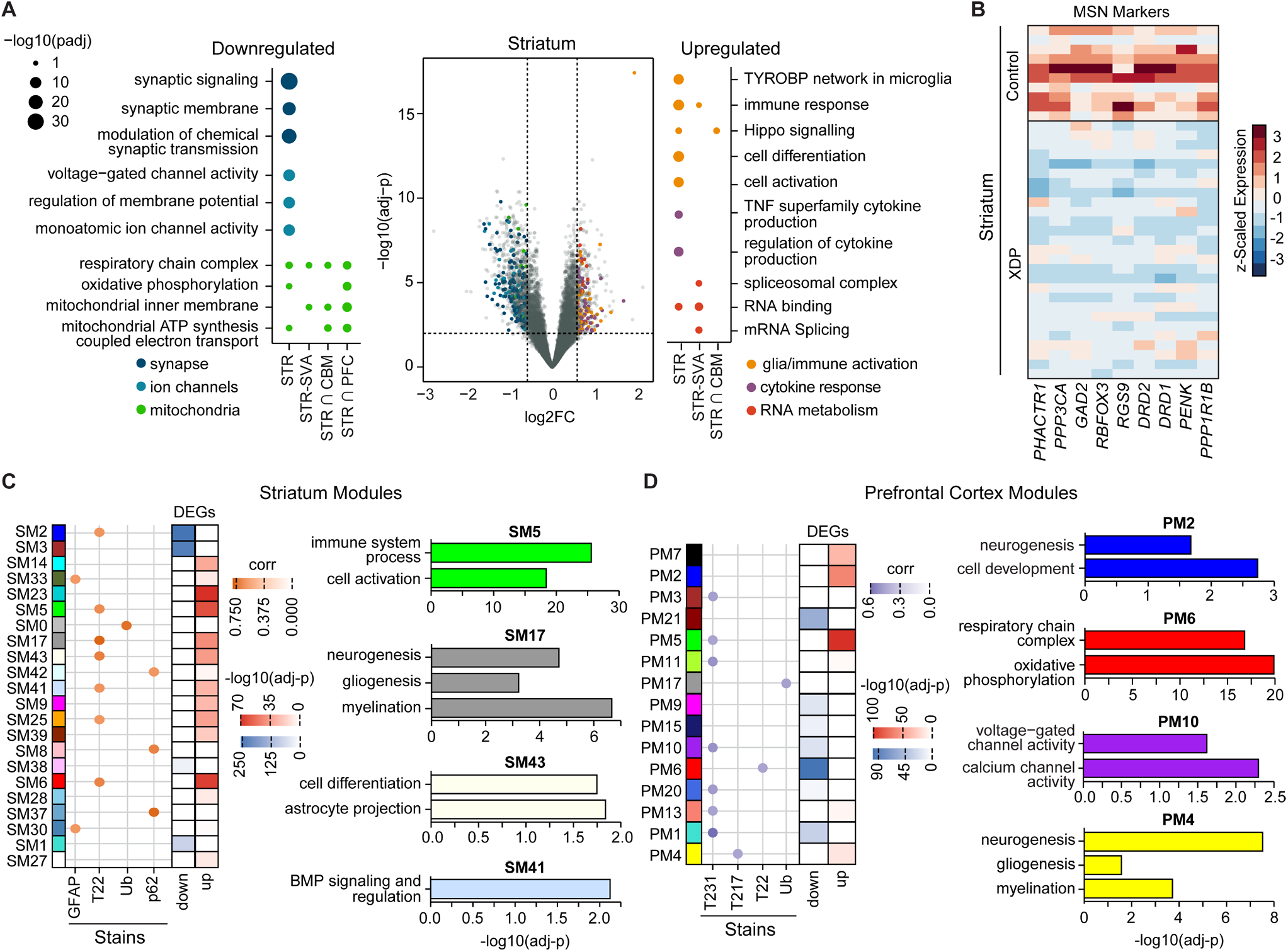
XDP brains display transcriptomic signatures of neurodegeneration, tau aggregation, and neuroinflammation. **A**, Differentially expressed genes (DEGs) were most profound in the striatum (STR), and are enriched for biological processes related to tau-mediated neurotoxicity (downregulated genes) and neuroinflammation (upregulated genes). These enrichments are also observed in STR DEGs overlapping with signatures from the cerebellum (CBM) and prefrontal cortex (PFC), as well as in a subset of STR DEGs that correlate with SVA (CCCTCT)_n_ repeat length (STR-SVA). A set of upregulated STR-SVA DEGs are enriched for RNA splicing ontologies. **B**, Canonical medium spiny neuron (MSN) markers are markedly downregulated in XDP STR. **C-D**, Eigengenes from gene co-expression modules identified from XDP STR (**C**) and PFC (**D**) correlate with phospho-tau (p-tau231, p-tau217) immunoreactivity, soluble misfolded tau (T22), 4R tau levels, and ubiquitin (Ub)/p62 staining. DEGs from these brain regions are distributed across these modules and converge on dysregulated biological processes in tau-affected regions, as supported by module preservation and functional enrichment analyses.

We performed functional enrichment analysis of striatal DEGs shared with other brain regions exhibiting tau pathology (**Figure 4A**). In parallel, we regressed normalized gene expression against (CCCTCT)_n_ repeat length to identify transcripts with expression levels that were associated with higher repeat lengths (**Table S4**). Ontologies related to mitochondrial function, including oxidative phosphorylation and the respiratory chain, were significantly enriched among downregulated striatal DEGs (adj-p=4.0e-03 for respiratory chain complex; all enriched terms adj-p<0.05, **Figure 4A, Table S5**) and were prominent in genes showing stronger downregulation with increasing repeat length. Mitochondrial pathways were also specifically enriched among downregulated striatal DEGs shared with prefrontal cortex and cerebellum, indicating a convergent transcriptional signature across tau-affected regions.

Downregulated striatal DEGs displayed pronounced enrichment for genes involved in neuronal synaptic function and voltage-gated ion channels, reflecting the loss of key components of neurotransmission in this region (**Figure 4A, Table S5**)^59^. Notably, these genes were strikingly concordant (95% overlap) with those that were reduced in degenerating MSNs in HD (p=1.0e-207)^60^, underscoring the shared transcriptional features of striatal degeneration between XDP and other repeat-mediated neurodegenerative disorders. Canonical markers of medium spiny neurons (MSNs) including *PPP1R1B, DRD1*, and *DRD2* were markedly downregulated (**Figure 4B**), consistent with MSN loss documented in prior neuropathological reports^26,27^. We found that *CALB1* was significantly decreased (log_2_FC=-1.5, adj-*p*=6.4e-05), while other markers of cholinergic interneurons were unchanged, suggesting a specific vulnerability of striatal MSNs. Functional enrichment among upregulated striatal DEGs implicated genes related to immune response, cell differentiation, and gliosis-associated pathways (all enrichments adj-*p*<0.01; **Figure 4A, Table S5**). A particular glial response pathway that was strongly enriched in upregulated striatal DEGs was TYROBP signaling (adj-*p*=9.6e-09), a microglial module that has been linked to tau pathology in frontotemporal dementia (FTD)^61^. The subset of upregulated striatal DEGs whose expression levels correlated directly with increasing (CCCTCT)_n_ repeat length was enriched for splicing- and RNA processing–related ontologies (adj-*p*=0.04) (**Figure 4A, Table S5**). This group included the transcript encoding the RNA-binding protein hnRNPC that regulates *MAPT* splicing^62^, as well as several ribosomal protein genes implicated in the tau interactome^63^. In addition, several members of the serine/arginine-rich protein family were dysregulated in XDP striatum including *SRSF2* (also known as SC35) that was previously shown to promote *MAPT* exon 10 inclusion^64,65^.

We hypothesized that transcriptional changes in the cortex might reveal disease-associated mechanisms that were less confounded by cell death. We thus compared co-expression modules derived from the striatum and prefrontal cortex and identified functional enrichments in convergent/preserved modules, focusing on gene sets whose expression profiles correlated with phospho- and/or misfolded tau immunostaining (adj-*p*<0.10; **Figure 4C-D and S9D-E, Table S6-S7**). This analysis revealed downregulated modules from the prefrontal cortex that were highly enriched for oxidative phosphorylation (PM6, adj-*p*=1.3e-20), respiratory-chain (PM6, adj-*p*=5.42e-15), and modestly associated with voltage-gated-channel activity (PM10, adj-*p*=0.02) (**Figure 4C, Table S8**), all processes that were also observed in downregulated striatal DEGs (**Figure 4A, Table S5**). Among these, PM6 genes overlapped a set of mitochondrial factors previously shown to promote tau aggregation in neuronal modifier screens^66,67^. Modules upregulated in the prefrontal cortex, including PM2, PM4, and PM13, were enriched for neurodevelopmental and glial-differentiation pathways that encompassed neurogenesis, gliogenesis, and myelination-related ontology terms (**Figures 4D and S9E, Table S8**). Of these gene sets, PM4 was among the most strongly preserved dysregulated cortical modules, corresponded to striatal module SM17 (**Figure 4C-D, Tables S8-S9**), and overlapped with biological processes previously reported in the prefrontal cortex of P301S tau transgenic mice^68^. Additional convergent modules from both regions (PM5 and SM41) were enriched for bone morphogenetic protein (BMP) signaling, identifying a potentially actionable tau-related pathway (**Figures 4C and S9E, Tables S8-S9**)^69^. *MAPT* itself was contained within a convergent prefrontal module (PM11) whose eigengene correlated with p-tau231 immunostaining (r=0.51, *p*=1.0e-03) (**Figure 4D, Tables S6-S7**).

We also investigated the correlation of coexpression modules with molecular (i.e., levels of *TAF1* intron 32 retention) and pathologic features (i.e., GFAP, ubiquitin, or p62 staining) (**Figure 4C-D, Table S7**). Aberrant splicing in *TAF1* intron 32 is the most consistent and robust transcriptomic signature of XDP in neural cell lines and represents one mechanism by which the SVA can induce TAF1 dysfunction^18,70^. One module, PM7, exhibited eigengene correlations with p-tau231 (r=0.34, *p*=0.03) as well as *TAF1* intron 32 retention (r=0.30, *p*=0.07, **Table S7**). This module overlapped with AD-associated cortical modules related to tau accumulation (adj-*p*=8.6e-11)^71^ and contained genes encoding members of the eIF4 complex (*EIF4B, EIF4G2, EIF4H, EIF4A2*) that interact with tau (**Figure 4D, Table S6**)^63,72^.

Together, these findings pointed to a consistent intersection between the XDP transcriptomic landscape and disease mechanisms in other tau-related disorders, motivating further investigation of *MAPT* regulation and tau isoform imbalance in XDP brains.

### Increased MAPT mRNA expression and pathogenic imbalance of tau isoforms in the XDP striatum

Building on transcriptomic findings of altered expression of factors known to regulate *MAPT* splicing, we quantified the expression of *MAPT*-associated transcripts in XDP brain tissues. Alternative splicing of *MAPT* generates six distinct tau isoforms, classified as four-repeat (4R) or three-repeat (3R) depending on the inclusion of an exon 10 that encodes one of the protein’s microtubule-binding repeats. In the adult human brain, 4R and 3R isoforms are normally expressed at comparable levels^73^, and both genetic^7–10^ and functional^13,74–77^ studies suggest that imbalances in this ratio can drive neurodegeneration and define specific tauopathies. In XDP brains, we found a significant increase in *MAPT* expression in the striatum (log_2_FC=0.33, adj-*p*=2.0e-04), as well as nominal increases in the prefrontal cortex and cerebellum (log_2_FC<0.20, adj-*p*<0.05) (**Figure 5A**). *MAPT* splicing analysis in XDP brains revealed increased inclusion of exon 10 compared to controls, measured as percent spliced-in (PSI), that was specific to the striatum (*p*=1.8e-02) (**Figure 5B**), and positively correlated with (CCCTCT)_n_ repeat length (r=0.52, *p*=9.0e-03) (**Figure 5C**). We further validated these findings by staining XDP (n=20) and control (n=5) striatal tissue with RD4 and RD3 antibodies, which have been previously validated for the isoform-specific detection of 4R and 3R tau, respectively^58,78^. We confirmed the specificity of these antibodies by immunofluorescence staining of Pick’s disease (primary 3R tauopathy), AD (mixed 3R/4R tauopathy), and progressive supranuclear palsy (primary 4R tauopathy) tissues, which showed the expected isoform-selective patterns (**Figure 5D**). From these analyses, XDP striatum again showed a significant increase in 4R tau levels relative to control tissue (*p*=2.4e-03) without a corresponding change in 3R tau (**Figure 5E-F, Table S2**).

**Figure 5.**
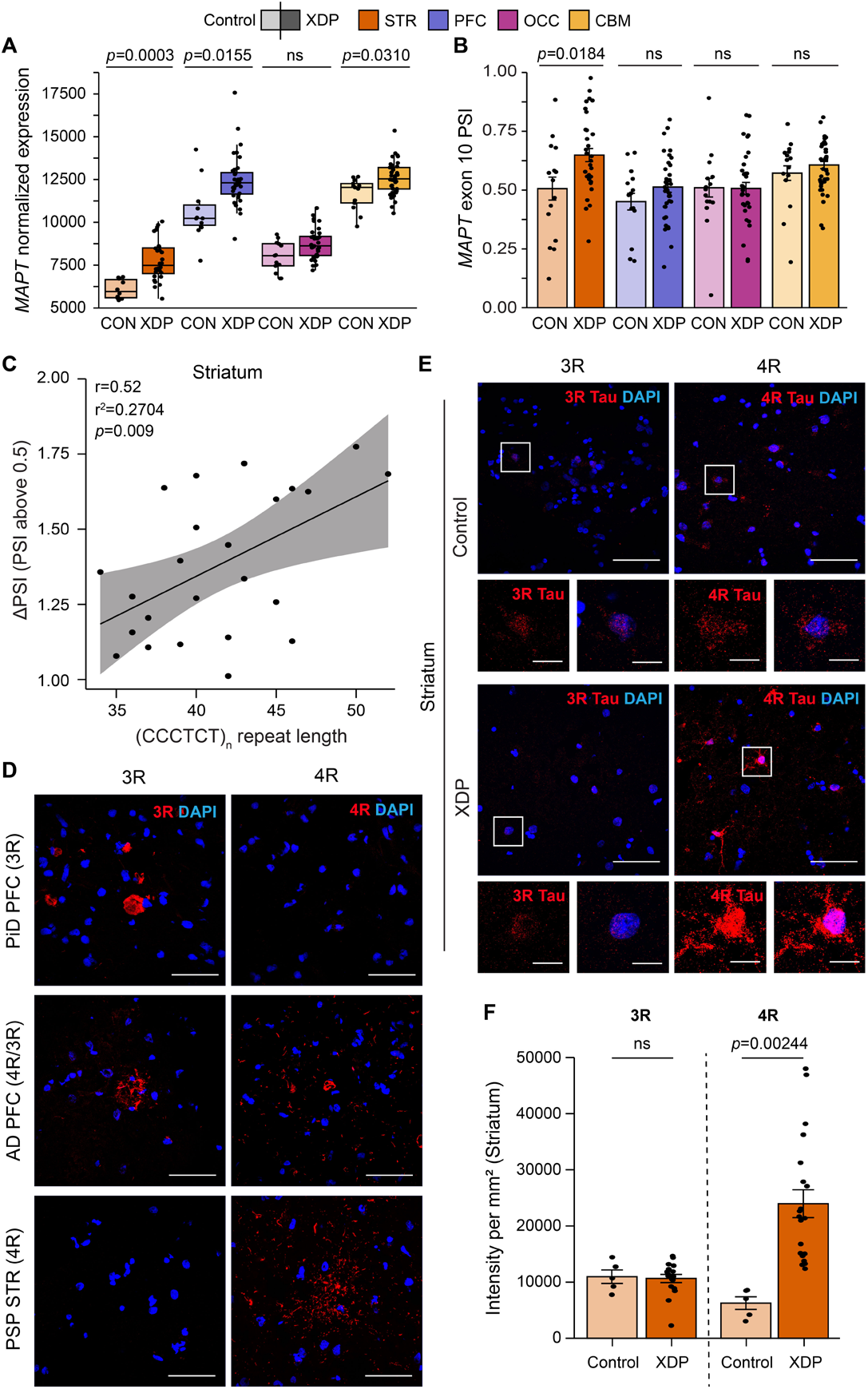
Increased *MAPT* expression and elevated 4R/3R tau isoform ratios in XDP striatum. **A**, *MAPT* mRNA expression is increased in the striatum (STR), cerebellum (CBM), and prefrontal cortex (PFC) in XDP, with no change in the occipital cortex (OCC). **B**, *MAPT* exon 10 inclusion (percent spliced-in, PSI) is elevated exclusively in XDP striatum. **C**, ΔPSI above the physiological *MAPT* exon 10 inclusion baseline (0.5) positively correlates with (CCCTCT)_n_ repeat length. **D**, Representative confocal images of 3R and 4R tau staining in Pick’s disease (PiD) prefrontal cortex (PFC; 3R tauopathy), Alzheimer’s disease (AD) prefrontal cortex (PFC; 3R/4R tauopathy), and progressive supranuclear palsy (PSP) striatum (STR; 4R tauopathy), validating isoform-specific antibody reactivity. **E**, Representative confocal images of control (n=5) and XDP (n=20) striatum stained for 3R and 4R tau. Insets show higher-magnification views of boxed regions. **F**, Quantification of tau isoforms shows increased 4R tau intensity in XDP striatum (*p*=0.00244), with no change in 3R tau. Data are mean ± s.e.m.; each point represents one case. Statistical significance is indicated in panels. Scale bars, 50 μm (overview images) and 10 μm (insets).

Because *MAPT* haplotypes can influence *MAPT* transcription, alternative splicing, and susceptibility to tauopathies^79–82^, we asked whether haplotype differences might account for the changes in tau expression and splicing observed in XDP. The *MAPT* H1 haplotype, which is associated with increased 4R tau expression and elevated risk for PSP^82^, was identified in all XDP and control subjects (n=68) by SNP genotyping, with all individuals being homozygous for the H1 haplotype (**Table S1**). Thus, the observed differences in *MAPT* expression and tau isoform ratios in the brain were not attributable to genetic variation in *MAPT* among tissue donors.

### MAPT dysregulation and 4R tau accumulation are associated with astrocyte reactivity and changes in cellular composition in the XDP striatum

Imbalances of tau isoform ratios are neurotoxic^74,76,77,83^ and linked to neuroinflammatory responses^75^. To determine whether shifts in cellular composition occur in XDP brains, we performed cell-type deconvolution of bulk RNA-seq profiles. In the striatum, we observed reduced neuronal cell populations consistent with MSN loss (**Figure 6A**), accompanied by increased proportions of astrocytes (*p*=0.03), microglia (*p*=1.9e-04), and oligodendrocytes (*p*=4.1e-05), as well as upregulation of *GFAP*, a canonical marker of astrocytic reactivity (log_2_FC=1.01, adj-*p*=3.7e-03). Recapitulating these cell-type inferences from bulk RNA-Seq, immunostaining confirmed extensive reactive astrogliosis in striatal tissue (**Figure 6B-C**). Although the prefrontal cortex did not exhibit major shifts in cell proportions and showed no significant alterations in *GFAP* expression, we did observe a nominal increase in the proportion of oligodendrocytes (*p*=5.8e-04; **Figure 6A**) and a modest reduction in the neuronal cell fraction, which aligns with prior neuroimaging evidence of cortical involvement in XDP^28,29^.

**Figure 6.**
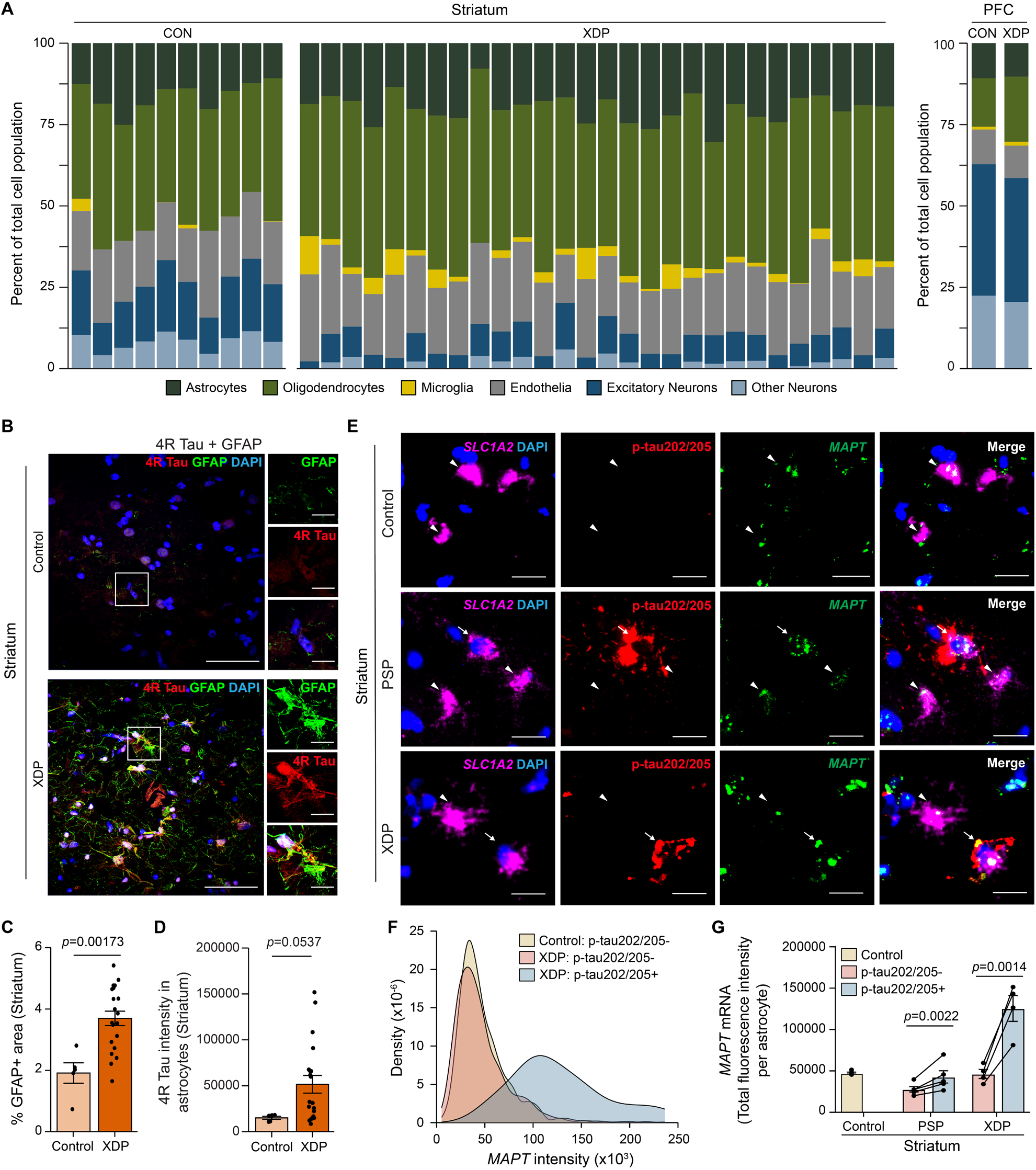
*MAPT* dysregulation and 4R tau accumulation are associated with astrocyte reactivity and altered cellular composition in the XDP striatum. **A**, Cell-type deconvolution of bulk RNA-seq data shows altered proportions of neuronal and glial populations in the striatum of XDP compared to control, with increased oligodendrocyte and astrocyte fractions and reduced neuronal proportions. A modest reduction in neuronal fractions is also observed in the prefrontal cortex (PFC). Deconvolution is presented per-brain in striatum to show the variation, and grouped in PFC (across n=36 XDP and 11 controls). **B**, Representative confocal images of control and XDP striatum co-stained for 4R tau and GFAP, showing increased astrocytic 4R tau accumulation in XDP. Insets show higher-magnification views of boxed regions. **C**, Quantification of GFAP-positive area demonstrates increased astrocyte reactivity in XDP striatum. **D**, Quantification of 4R tau intensity within astrocytes shows increased 4R tau accumulation in XDP striatum. **E**, RNAscope™ fluorescence *in situ* hybridization (FISH) for *SLC1A2* (astrocyte marker) and *MAPT* mRNAs combined with p-tau202/205 (AT8) immunostaining in control, progressive supranuclear palsy (PSP), and XDP striatum. Arrowheads indicate astrocytes co-expressing *MAPT* and p-tau202/205 (AT8). **F**, Distribution of *MAPT* transcript intensity in astrocytes stratified by p-tau202/205 status shows higher *MAPT* levels in p-tau202/205-positive astrocytes in XDP striatum. **G**, Quantification of the mean *MAPT* mRNA fluorescence intensity per astrocyte shows consistently increased *MAPT* levels in p-tau202/205-positive astrocytes compared to p-tau202/205-negative astrocytes in PSP and XDP striatum. Data are mean ± s.e.m.; each point represents one case. Statistical significance is indicated in panels. Scale bars, 50 μm (overview images) and 10 μm (insets).

Thus, both the transcriptomic and histopathology analyses revealed substantial astrogliosis in XDP brains. Astrocytes are particularly sensitive to alterations in the 4R/3R tau ratio^75,84^, and increased 4R tau drives a neurotoxic reactive phenotype accompanied by impaired homeostatic functions^95^. Using co-labeling with RD4 and GFAP antibodies, we observed elevated 4R tau within XDP striatal astrocytes compared to control (*p*=0.05), supporting a link between tau isoform imbalance and astrocyte reactivity in XDP (**Figure 6B,D**).

We next asked whether tau accumulation within astrocytes reflected uptake of protein released by neighboring cells or resulted, at least in part, from increased *MAPT* transcription. Indeed, although *MAPT* expression is predominantly neuronal, astrocytes express detectable levels of *MAPT* mRNA under physiological conditions^64,65^ with further elevations detected in astrocytes bearing tau inclusions^66^. RNA fluorescence *in situ* hybridization (FISH) for *MAPT* and the astrocytic marker, *SLC1A2*, combined with immunofluorescence for p-tau202/205 (AT8), was performed to compare *MAPT* transcript levels in astrocytes with or without tau aggregates (n=5 controls, n=4 XDP). Striatal tissue from patients with PSP (n=5) served as positive controls (**Figure 6E**). In both PSP (*p*=2.3e-03) and XDP (*p*=1.45e-03), p-tau202/205-positive astrocytes displayed significantly higher *MAPT* mRNA levels than p-tau202/205-negative astrocytes from the same donor (**Figure 6E-G, Table S10**), indicating that *MAPT* expression is increased in striatal astrocytes harboring pathological tau aggregates in XDP. Together, these findings suggest that dysregulated *MAPT* expression and splicing may drive changes in cellular composition in XDP brains, in part through astrocyte-mediated inflammation.

### Plasma NfL, GFAP, and p-tau181 are elevated in XDP plasma

Having identified tau-related molecular and cellular disturbances in XDP brain tissue, we next asked whether corresponding signatures could be detected in peripheral biofluids from living patients with XDP (n=119) versus their SVA-negative, healthy male relatives (n=29). Plasma phosphorylated tau (p-tau) is among the most sensitive biomarkers for early AD^85–87^, yet similarly effective fluid biomarkers for other tauopathies have been limited^88^. We therefore assayed p-tau181 and total tau (tTau), as well as neurofilament light chain (NfL) as a general sensor of neurodegeneration and three markers of glial activation or inflammation (GFAP, fractalkine, and YKL-40) in plasma collected from these individuals (**Figures 7A-G and S10**). To account for the known effects of aging on plasma biomarker concentrations, group comparisons were performed using age-adjusted linear regression models on log-transformed biomarker values. NfL (*p*=1.5e-03; area under the curve (AUC=0.722) and GFAP (*p*=6.3e-04; AUC=0.693) were significantly elevated in XDP plasma compared to controls and showed comparable discriminatory performance (**Figure 7A-B, G**). Among tau-related measures, p-tau181 was significantly higher in XDP (*p*=1.9e-02; AUC=0.649), while total tau (tTau) did not significantly differ between groups (*p*=0.11) (**Figure 7C,E,G**). Fractalkine did not differ significantly between groups (*p*=0.28) and YKL-40 levels were higher in controls (*p*=1.8e-02) (**Figure 7D,F,G**). Together, these findings identify NfL, GFAP, and p-tau181 as candidate plasma biomarkers of neurodegeneration and tau pathology in XDP.

**Figure 7.**
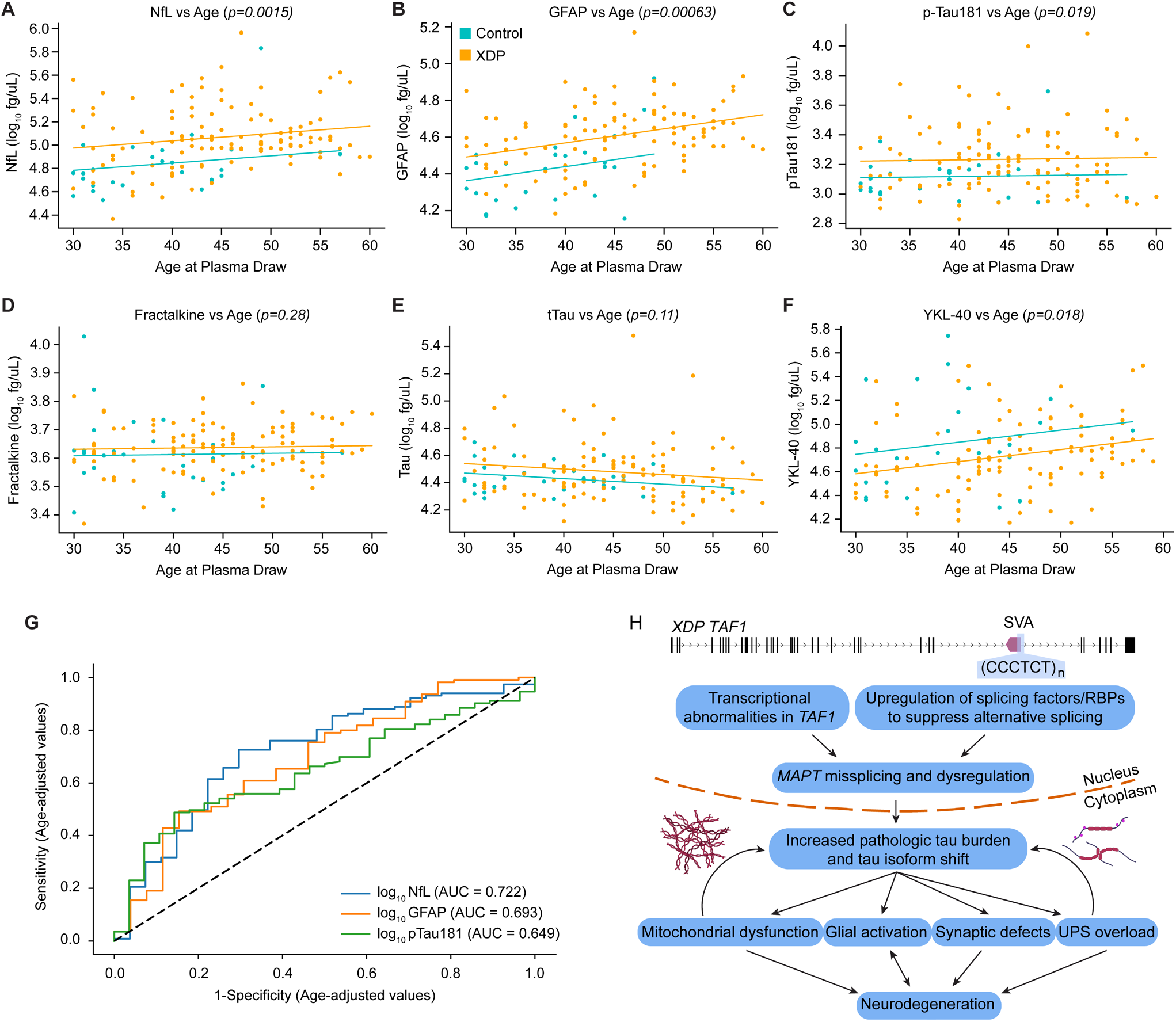
Age-associated plasma biomarker changes and a proposed pathogenic cascade linking TAF1 dysfunction to tau pathology in XDP. **(A-F)** Associations between age at plasma collection and plasma concentrations of neurofilament light chain (NfL) (**A**), glial fibrillary acidic protein (GFAP) (**B**), phosphorylated tau-181 (p-tau181) (**C**), fractalkine (**D**), total tau (tTau) (**E**), and YKL-40 (**F**) in control (n=29) and X-linked dystonia-parkinsonism (XDP; n=119) participants from the age-restricted cohort (30-60 years at plasma collection). Biomarker values were log-transformed prior to analysis to account for right-skewed distributions. Lines indicate linear regression fits for each group, and the *p* values shown in each panel reflect the significance of differences between XDP and control participants after adjustment for age. (**G**) Receiver operating characteristic (ROC) analyses based on age-adjusted residuals derived from rank-based ANCOVA demonstrating the discriminatory performance of plasma biomarkers for distinguishing XDP from controls. NfL exhibited the highest performance (AUC=0.722), followed by GFAP (AUC=0.693) and p-tau181 (AUC=0.649). (**H**) Schematic model of the proposed pathogenic cascade in XDP. A polymorphic (CCCTCT)_n_ repeat within an intronic SINE-VNTR-Alu (SVA) retrotransposon in TAF1 results in transcriptional abnormalities and altered expression of splicing factors and RNA-binding proteins (RBPs), leading to MAPT missplicing and dysregulation. These changes promote increased pathological tau burden and tau isoform imbalance, which may contribute to mitochondrial dysfunction, glial activation, synaptic defects, and ubiquitin-proteasome system (UPS) overload, ultimately converging on neurodegeneration. Nuclear and cytoplasmic compartments are indicated for orientation. Each point represents one participant.

## DISCUSSION

The 4R tauopathies comprise a heterogeneous group of neurodegenerative disorders characterized by the aggregation and spreading of 4R tau, resulting in characteristic clinical features that are mainly classified as movement and/or cognitive disorders^89^. Through comprehensive neuropathological, biochemical, and transcriptomic analyses of a large collection of XDP brains, we found multiple lines of evidence indicating that XDP shares key pathological characteristics with the 4R tauopathies. Ubiquitinated and p62-immunoreactive aggregates containing pathological tau were detected throughout the XDP striatum, prefrontal cortex, and cerebellum. In addition, abundant misfolded, potentially oligomeric forms of tau, which are thought to be the most neurotoxic tau species in other disorders^42–44^, accumulated in the striatum at levels that significantly correlated with the number of repeats in the XDP causal mutation. Molecular analyses consistently linked dysregulated gene sets in XDP brain to known tau-related cascades, including enrichment of DEGs related to mitochondrial dysfunction, a pathway that potently promotes tau oligomerization through the proteasomal misprocessing of tau^66,67^. Finally, we detected a pathogenic skewing of tau isoform ratios^7–10,74–76,83^ in favor of the longer, more aggregation-prone^90–93^ 4R tau isoform, an upstream driver of tau hyperphosphorylation and oligomerization^74^. Collectively, these findings position XDP as a distinct genetic model for dissecting how mobile element insertions and repeat expansions may initiate tau-driven neurotoxicity, with potential relevance to related neurodegenerative diseases.

A key feature of the pathology in XDP brains was the accumulation of misfolded tau largely within astrocytes rather than neurons in the striatum. We further observed increased 4R tau protein in astrocytes, as well as elevated *MAPT* mRNA levels in those bearing tau aggregates, indicating that tau accumulation in XDP astrocytes may involve both intrinsic upregulation and uptake. The origin and pathogenic role of astrocytic tau inclusions have been debated across tauopathies; while early studies suggested that they largely reflect secondary accumulation derived from neighboring tau-affected neurons^94,95^, recent evidence indicates that astroglial tau can arise through cell-intrinsic mechanisms and directly induce neurotoxicity^75,96^. Astrocytic tau pathology reportedly precedes neuronal tangle formation in the striatum of PSP^97^ and corticobasal degeneration (CBD)^98^, and similar tau-immunoreactive astrocytes within the substantia nigra have been observed in nigral tau astrogliopathy (NITAG)^99^. Consistent with these observations in human tissue, transgenic mice overexpressing human tau in astrocytes develop inclusions typical of 4R tauopathies^96^. Our findings paralleled recent transcriptomic and neuropathological studies showing *MAPT* upregulation in tau-laden astrocytes^100–104^, supporting a model in which astrocytic tau contributes directly to disease propagation and neurodegeneration in XDP.

To investigate the molecular mechanisms underlying tau pathology and regional vulnerability in XDP, we analyzed RNA-seq data from multiple brain regions, including the prefrontal cortex and striatum. Given the distinct states of pathological tau detected in these regions, this analysis offered a systems-level view of transcriptional changes that occur in brain regions with and without overt neuronal loss. Co-expression network analyses revealed gene modules linking tau propagation, *MAPT* expression, and *TAF1* splicing abnormalities characteristic of XDP. These modules overlapped with pathways previously implicated in tau aggregation and clearance, and the regional comparisons indicated that the molecular perturbations driving tau accumulation in the striatum were also present in cortex. Notably, the results further suggested that *MAPT* dysregulation in XDP may be influenced by the causal *TAF1* mutation. Indeed, correlations between the pathogenic (CCCTCT)_n_ repeat length, exon 10 inclusion, and misfolded tau levels are consistent with a quantitative relationship linking the repeat insertion to skewed 4R/3R isoform ratios and tau accumulation. Evidence from other neurodegenerative diseases supports this connection: HD exhibits features of TAF1 loss of function^105^ alongside pathological tau aggregation^106–108^, and a shift toward increased 4R/3R tau isoform ratios^106^. Together with recent findings that RAN-translated dipeptides derived from an SVA insertion at the CASP8 locus are linked to increased tau phosphorylation and aggregation^109,110^, these observations suggest multiple molecular genetic mechanisms by which repeat expansion diseases can harbor tau pathology.

**Figure 7H** outlines a proposed sequence of events based on these observations in striatum and cortex. In this model, the SVA retroelement insertion in *TAF1* contributes mechanistically to altered *MAPT* splicing, resulting in skewed tau isoform ratios that promote pathogenic tau deposition; this accumulation in turn may lead to mitochondrial disturbances that culminate in synaptic dysfunction and ultimately neurodegeneration. Further supporting this model, we detected increased expression of the splicing factors *HNRNPC* and *SRSF2*, both of which have been shown to promote exon 10 inclusion in MAPT pre-mRNA^62,65^ and protect the transcriptome by binding to transposable elements^111^. Notably, *HNRNPC* expression also correlated with repeat length in XDP striatum, and publicly available *TAF1* ChIP-seq datasets identify binding sites for TAF1 in the *MAPT* promoter region (encodeproject.org/experiments/ ENCSR000BTX/). Together, these findings suggest that the *TAF1* SVA insertion may perturb both transcriptional and splicing regulation of *MAPT*, fostering 4R tau accumulation and associated proteostatic stress. Future studies in XDP model systems will be essential to directly interrogate these potential regulatory interactions and determine how SVA-driven TAF1 protein dysfunction contributes to tau dysregulation.

The biochemical and functional properties of the tau species in striatum and cortex remain unclear as the soluble misfolded tau extracted from XDP brains did not exhibit measurable seeding activity in multiple biosensor assays. Notably, the finding that p-tau202/205-positive astrocytes in XDP exhibit elevated *MAPT* mRNA expression raises the possibility that seed-competent tau species may preferentially accumulate within astrocytes and are consequently underrepresented in bulk PBS-soluble homogenates. It is also possible that XDP-associated tau conformations are not efficiently detected by current biosensor systems, reflecting a conformational seeding barrier that limits cross-seeding with the reporter constructs used, thereby necessitating the development of biosensors tailored to XDP tau species^57,77^. Notably, emerging evidence from genetically defined tau models suggests that distinct mutations can differentially promote either tau hyperphosphorylation or seed-competent aggregation, representing separable early pathogenic trajectories^77,112,113^. In this context, XDP may preferentially engage a phosphorylation-dominant pathway, characterized by robust tau hyperphosphorylation and isoform imbalance without the early generation of readily detectable seed-competent species^77,113^. Together, these considerations raise the possibility that the lack of measurable seeding bioactivity reflects both technical limitations and intrinsic biological properties of XDP tau assemblies.

Although elevated tau levels were also detected in the XDP cerebellum, the observed pattern reflected a distinct pathological profile. In contrast to the striatum and cortex, cerebellar tau in XDP brains was predominantly localized within corpora amylacea with little to no evidence of hyperphosphorylation. The cellular origin and functional significance of corpora amylacea remain uncertain, yet increasing evidence suggests that their formation may represent a compensatory response to proteostatic or metabolic stress^40,41^. In AD models, the density of PAS granule, the murine counterparts of corpora amylacea, increases alongside early tau accumulation at early Braak stages yet declines at late stages, consistent with an adaptive but ultimately overwhelmed protective mechanism^41^. In XDP, the presence of tau-immunoreactive corpora amylacea in the cerebellar molecular layer, despite the absence of overt tau aggregates, supports the notion that their formation may sequester potentially early-stage toxic tau species or related metabolic by-products^40,41^. This pattern parallels proteomic data from transgenic tauopathy models showing upregulation of carbohydrate metabolism and protein catabolism pathways in the cerebellum^114^, processes linked to corpora amylacea formation and cellular resilience. Together, these observations raise the possibility that corpora amylacea formation may contribute to the relative sparing of the cerebellum in XDP and represent an endogenous neuroprotective response to tau stress.

While the detection of phosphorylated tau proteoforms in biofluids has been a turning point in the diagnosis of AD, their utility as diagnostic biomarkers for primary tauopathies is currently limited. Among these tau species, p-tau181 is a marker of synapses and neurites in healthy aging and AD brains^37,38^, and its concentration in plasma is significantly increased in patients with different neurodegenerative diseases^115–117^ and predicts cognitive decline in AD^38^. Intriguingly, there is evidence that plasma p-tau181 levels in AD may be increased only in amyloid-β-positive, cognitively impaired individuals with concomitant increases in plasma GFAP, underscoring the contribution of astrocyte reactivity as a critical mediator of tau pathology^118^. Although plasma GFAP levels are increased in a variety of neurological conditions^119^, they appear to correlate not only with cognitive decline but also tau pathology and reactive astrocytosis in post-mortem brain tissue from patients with AD^120,121^. Our findings that p-tau181 and GFAP are increased in plasma of patients with XDP similarly corroborate the increased accumulation of these proteins in brains from a separate cohort of XDP tissue donors. The fact that GFAP discriminated patients with XDP from controls with comparable efficacy as NfL raises the possibility that it could serve as a non-invasive biomarker in XDP. Future longitudinal studies with quantitative clinical assessments in XDP cohorts will be essential to determine how plasma GFAP and/or tau proteoforms correlate with specific clinical features and/or disease progression to validate their potential utility as biofluid-based endpoints in future clinical trials.

Collectively, our work integrating neuropathology, transcriptomics, and plasma protein profiling converges on defective tau proteostasis and astrocyte dysfunction as key contributors to neurodegeneration in XDP. In AD and other tauopathies, there has been considerable progress in lowering total tau levels^122,123^ and/or normalizing isoform ratios^74,75,124^ using multiple strategies including antisense oligonucleotides, immunotherapies, and small-molecule drugs^125^. Our study suggests that such approaches may eventually be applicable to XDP as well. Although most tauopathies arise sporadically, the identification of genetic variation linked to tau accumulation provides a unique opportunity to dissect the molecular and cellular mechanisms underlying tau dysregulation. In this context, XDP offers a genetically defined system for interrogating how a mobile element insertion perturbs tau homeostasis in patient cohorts alongside corresponding cellular and animal models. Additionally, these findings highlight how investigation of this rare hereditary neurodegenerative disorder can uncover potential links between a pathogenic repeat within an SVA insertion and tau-driven neurodegeneration.

## Data Availability

RNA-seq data have been deposited in dbGaP (Project phs001525.v2.p1).
Scripts used for preprocessing and analysis are available at github.com/
talkowski-lab/XDP_postmortembrains.

https://github.com/talkowski-lab/XDP_postmortembrains

## RESOURCE AVAILABILITY

RNA-seq data have been deposited in dbGaP (Project phs001525.v2.p1). Scripts used for preprocessing and analysis are available at github.com/talkowski-lab/XDP_postmortembrains.

## ACKNOWLEDGEMENTS

The authors extend their deepest gratitude to the patients with XDP and their family members for their invaluable participation and generosity in providing biospecimens that made this research possible. The authors also thank Dr. Amy Alessi, Ms. Trisha Multhaupt-Buell, and Ms. Gabrielle Aldykiewicz for expert administrative assistance in coordinating ethics approvals and biospecimen distribution. We thank all members of the Talkowski, Bragg, and Lagier-Tourenne labs for helpful discussions and support. This work was supported by the Collaborative Center for X-Linked Dystonia-Parkinsonism and by the National Institutes of Health (NIH)/National Institute of Neurological Disorders and Stroke (NINDS) grant R01NS102423 (M.E.T., D.C.B., C.L.-T.). M.E.T. is supported by NIH grant R01HD096326. D.G. is supported by the NIH/NINDS grant R00NS118109. N.R. is supported by a Byrne Family and Judith & Pape Adams Fellowship and a Fred and Gilda Slifka Neuroscience Transformative award. M.N. is supported by the Association for Frontotemporal Degeneration Holloway Postdoctoral Fellowship, The Cullen Education and Research Foundation, Hop On a Cure, and the Muscular Dystrophy Association. N.Q., R.E.B., and B.T.H. are supported by the Rainwater Charitable Foundation, the Cure Alzheimer’s Fund, and the Freedom Together Foundation. C.L.-T. is the recipient of the Araminta Broch-Healey Endowed Chair in ALS.

## AUTHOR CONTRIBUTIONS

C.J.R., A.D., E.B.P., M.E.T., D.C.B., and C.L.-T. conceived and designed the study. C.J.R., A.D., E.B.P., J.H., M.G.M., N.A.B.-V., H.-D.T., N.Q., A.M.d.G., N.S.-C., Y.T., N.R., B.W., A.H., R.Z.M., L.M., H.W., Y.Y.R., G.G., G.A.F., C.-Z.L., G.C., M.N., J.L., K.O., B.J., M.H., T.P., and P.K. performed the major experimental work. C.J.R., A.D., E.B.P., R.Y., D.G., S.R., S.E., J.M., and C.A.V. performed data analysis and visualization. E.N., J.H., M.G.M., C.F.-C., M.S.V.-A., G.P.A.L., N.G.G.-B., M.S., E.L.M., M.A.C.A., C.C.E.D., C.G., and N.S. coordinated and contributed to biospecimen collection. G.S.-V., M.W.A., S.A., B.J.W., R.E.B., M.I.D., B.T.H., and L.J.O. provided scientific input, access to resources, and/or supervision of specific experiments. C.J.R., A.D., E.B.P., C.A.V., M.E.T., D.C.B., and C.L.-T. wrote and revised the manuscript. M.E.T., D.C.B., and C.L.-T. supervised the study overall. All authors discussed the results and approved the final version of the manuscript.

## DECLARATION OF INTERESTS

C.L.-T. is a member of the Scientific Advisory Board and/or have received consulting fees from Arbor Biotechnologies Inc, AUTTX LLC, Dewpoint Therapeutics Inc, Libra Therapeutics, Mitsubishi Tanabe Pharma Corporation, Sanofi S.A., SOLA Biosciences, Nereid Therapeutics Inc, the Milken Institute, the Muscular Dystrophy Association, the Packard Center for ALS Research at Johns Hopkins, and the Northeast Amyotrophic Lateral Sclerosis Consortium. N.S. has received consulting fees from AbbVie Inc.

## MATERIALS AND METHODS

### Human biospecimens

All human studies were conducted in accordance with the Declaration of Helsinki and approved by the Institutional Review Boards (IRBs) of Massachusetts General Brigham (MGB; Boston, MA, USA) and the following institutions as indicated: Makati Medical Center (Makati, National Capital Region, Philippines), West Visayas State University (Iloilo City, Iloilo, Philippines), and José R. Reyes Memorial Medical Center (JRRMMC; Manila, Philippines). Written informed consent was obtained from all living participants, and post-mortem brain tissue specimens distributed for this study by the XDP Brain Bank maintained by the Collaborative Center for X-Linked Dystonia-Parkinsonism (CCXDP) were collected with informed consent from donors’ next of kin. The use of previously established iPSC lines was approved under MGH IRB protocols, and all procedures adhered to institutional and federal guidelines for research involving human biospecimens and derived cell lines.

#### Human post-mortem brain tissue

Post-mortem human brain samples were provided by the XDP Brain Tissue Bank maintained by CCXDP based at MGB. All procedures carried out by the CCXDP Bank related to engagement with tissue donors and family members, informed consent, tissue collection and processing, deidentified sample storage, and distribution are governed by protocols approved by Institutional Review Boards at MGB, Makati Medical Center, and West Visayas State University. Detailed descriptions of these methods, as well as quality control analyses of the tissue, have been recently reported^32^. Demographics of the samples used in the present study, as well as a matrix of which individuals were used for each experiment, are listed in **Table S1**.

#### Human induced pluripotent stem cell (iPSC) lines

Induced pluripotent stem cell (iPSC) lines used in this study included lines derived from (i) affected individuals with XDP; and (ii) unaffected control relatives confirmed to be negative for the causal SVA insertion; as well as (iii) CRISPR–Cas9–edited isogenic XDP lines in which the SVA element was excised (ΔSVA). Reprogramming of fibroblasts to iPSCs as well as neural conversion and differentiation to neural stem cells (NSCs) was previously reported^126^ along with CRISPR-editing to generate isogenic XDP control lines^18,70^.

#### Plasma from patients with XDP and unaffected control individuals

Peripheral blood was collected from male participants recruited through MGH and JRRMMC, as well as a regional clinic operated in Panay (Sunshine Care Foundation, Roxas City, Panay, Philippines). Plasma was isolated and processed as previously described^101^. Genomic DNA extracted from whole blood was used to confirm XDP genotype by PCR amplification of a 48-bp deletion haplotype marker^19,33^.

### Tissue staining and imaging

All staining and hybridization procedures were performed at room temperature unless otherwise specified.

#### Immunofluorescence staining of human brain tissue

Fresh-frozen human brain tissues were sectioned at 10 µm, mounted onto Superfrost™ Plus microscope slides (Fisher Scientific, 12-550-15), fixed with 10% neutral-buffered formalin (Millipore Sigma, HT501128-4L) for 10 min, and placed in Coverplate™ disposable immunostaining chambers (ThermoFisher Scientific, 72110017) and Epredia™ Shandon™ Sequenza™ immunostaining slide racks (ThermoFisher Scientific, 73310017). Sections were then permeabilized in 0.4% Triton X-100 in 1× PBS for 30 min, washed three times (5 min each) in 0.1% Triton X-100 in 1× PBS, blocked with 10% normal goat serum (Abcam, ab7481) in wash buffer for 1 h, and incubated overnight at 4 °C with primary antibodies (**Table S11**). Tissues were subsequently washed three times (10 min each) in wash buffer and incubated with secondary antibodies for 2 h (**Table S12**), followed by three additional washes (10 min each). Slides were then counterstained with 1× DAPI (ThermoFisher Scientific, D1306) for 15 min, treated with 0.1% Sudan Black B (Millipore Sigma, 4197-25-5) in 70% ethanol (Fisher Scientific, BP2818100) for 10 min to quench autofluorescence, cover-slipped using ProLong™ Glass Antifade Mountant with NucBlue™ Stain (ThermoFisher Scientific, P36981), and stored in the dark at 4 °C prior to imaging on a Zeiss LSM 900 confocal microscope with Airyscan 2.

#### Thioflavin S (ThioS) staining

Following completion of immunofluorescence staining and secondary antibody incubation, tissue sections were incubated in 0.05% ThioS solution (Sigma, T1892) for 8 min in the dark. Slides were briefly differentiated by immersion in 80% ethanol for 15 s and subsequently washed four times in distilled water for 5 min each. Sections were then processed for nuclear counterstaining, autofluorescence quenching, and mounting as described above.

#### Periodic acid–Schiff (PAS) staining

PAS staining was performed on human brain tissue sections to detect the polysaccharide-rich core of corpora amylacea using a commercial PAS stain kit (Abcam, ab150680) with minor modifications. Fresh-frozen human cerebellar sections (10 µm) were air-dried for 3-5 min and fixed overnight in 10% neutral-buffered formalin (Fisher Scientific, SF100-20). Following fixation, slides were rinsed with distilled water and incubated in PAS solution for 5-10 min. After four washes in distilled water, sections were incubated in Schiff’s reagent for 15-30 min and rinsed again in distilled water until the background cleared. Nuclei were counterstained with hematoxylin for 2-3 min, blued in bluing reagent for 30 s, and rinsed once more in distilled water. Slides were then incubated in Light Green solution for 2 min, washed briefly in distilled water, dehydrated through graded ethanol (70%, 95%, and 100%; two changes each, 10 dips per change), air-dried, and cover-slipped using mounting medium.

#### RNAscope fluorescence in situ hybridization

RNA fluorescence in situ hybridization was performed using the RNAscope™ Multiplex Fluorescent Assay v2 kit (ACD, 323100) following a previously published protocol^102^ with minor modifications. Fresh-frozen brain sections were immersed in 10% neutral-buffered formalin (Millipore Sigma, HT501128-4L) for 10 min at 4°C, then dehydrated in 50% ethanol for 5 min, 70% ethanol for 5 min, and twice in absolute ethanol (Fisher Scientific, BP2818100) for 5 min each. Slides were air-dried, and tissue boundaries were outlined using an ImmEdge hydrophobic barrier pen (ACD, 310018). Sections were treated with RNAscope™ hydrogen peroxide solution (ACD, 322335) for 10 min, followed by incubation with RNAscope™ Protease IV reagent (ACD, 322340) for 5 min. Slides were hybridized with target probes for 2 h at 40 °C in a HybEZ™ II hybridization oven (ACD, 321720).

All tissues were hybridized with RNAscope™ 3-plex Positive Control Probe-Hs (ACD, 320861) and RNAscope™ 3-plex Negative Control Probe (ACD, 320871) to assess RNA integrity. Slides with compromised RNA integrity were excluded from further analysis. Sections passing quality control were hybridized with RNAscope™ Probe-Hs-MAPT (ACD, 408991) and RNAscope™ Probe-Hs-SLC1A2-C3 (ACD, 444721-C3) to examine MAPT mRNA expression in astrocytes. Slides were stored overnight in 5× saline–sodium citrate buffer (Promega, V4261) before consecutive amplification steps were performed in the HybEZ™ II oven according to the manufacturer’s instructions. C1 probes were developed with the TSA Plus Cy3 detection kit (Akoya Biosciences, NEL744E001KT), and C3 probes were developed with the TSA Plus Fluorescein detection kit (Akoya Biosciences, NEL741001KT). C2 probes, were developed with the TSA Plus Cy5 detection kit (Akoya Biosciences, NEL745001KT) in some experiments as indicated.

Following probe development, sections were processed for immunofluorescence as described above, using the conformation-specific anti-p-tau202/205 (ThermoFisher Scientific, MN1020; 1:500) applied overnight at 4 °C, followed by donkey anti-mouse IgG (H+L), Alexa Fluor™ 568 secondary antibody (ThermoFisher Scientific, A10037; 1:1,000) for 2 h. Slides were then counterstained with 1× DAPI (ThermoFisher Scientific, D1306) for 15 min, treated with 1× TrueBlack™ lipofuscin autofluorescence quencher (Biotium, 23007) in 70% ethanol for 1 min to reduce autofluorescence, and cover-slipped using ProLong™ Glass Antifade Mountant with NucBlue™ Stain (ThermoFisher Scientific, P36981). Slides were stored in the dark at 4 °C until imaging on a Zeiss LSM 900 confocal microscope with Airyscan 2 or a NanoZoomer® S60 whole-slide scanner (Hamamatsu, C13210).

#### Quantification of imaging data and statistical analyses

Slides were digitized using a Nanozoomer® S60 whole slide scanner (Hamamatsu, C13210) at 40x resolution. Digital images were subsequently analyzed with QuPath^127^, an open-source software platform for bioimage analysis. For each brain region, up to 10 regions of interest (ROIs) were systematically selected for quantification and downstream analysis by an investigator blinded to genotype. Statistical analyses and plotting were performed using R v4.4.2.

For comparisons of total ubiquitin/p62-positive inclusions, a generalized linear model was used with age-at-death included as a covariate. For comparisons of total, misfolded, and phospho-tau species, within-individual variation across repeated measures of immunostaining intensity and immunoreactive area was modeled by including the individual as a random effect in generalized linear mixed models. Means across ROIs were used for plotting group-level comparisons between XDP and controls, and error bars represent the standard error of the mean (SEM). Correlations of (CCCTCT)_n_ repeat length, demographic variables, and tau measurements were assessed using Pearson correlation. Differences in *MAPT* transcript intensities measured via RNAScope between p-tau202/205-positive and -negative astrocytes were evaluated using multiple t-tests.

### Protein extraction

All procedures were carried out on ice or at 4 °C unless otherwise specified.

#### Total brain lysate and insoluble protein extraction for v2L P301S tau seeding assays

Frozen tissue samples (100 mg) were homogenized in 1× Tris-buffered saline (TBS) supplemented with a protease inhibitor cocktail (PIC) (Thermofisher Scientific, A32965). Protein concentrations were determined using the Pierce 660 nm Protein Assay and normalized to 1.6 mg/mL. Total lysates were used directly for tau seeding assays by adding 10 μg of total protein to v2L P301S biosensor cells.

To prepare the insoluble fraction, 200 μg of total lysate was ultracentrifuged at 186,000 × g for 1 h at 4 °C. The supernatant was discarded, and the pellet was resuspended in 50 μL of the same lysis buffer, followed by sonication. A 3 μL aliquot of the resulting suspension was added per well to v2L P301S biosensor cells for tau seeding assays.

#### PBS-soluble protein extraction for wild-type 3R/4R and 4R/4R tau seeding assays

PBS-soluble protein fractions from the prefrontal cortex and striatum were collected as previously described^44^. Briefly, 500 mg of prefrontal cortex and 50 mg of striatum were homogenized in 2.5 mL (5 v/w) of 1× PBS containing protease inhibitor cocktail (Cell Signaling Technology, 5871) using a 15 mL glass homogenizer (30 up-and-down strokes, 70% power) on ice with a Glas-Col® High-Speed Homogenizer (Cole-Parmer, 099C K54). Brain homogenates were transferred to 50 mL Falcon™ conical tubes (Fisher Scientific, 352070) and centrifuged at 10,000 g for 10 min. The resulting supernatants (PBS-soluble fraction) and pellets (PBS-insoluble fraction) were collected and stored at −80 °C until further use.

### Cell culture and functional assays

All cell culture procedures were conducted under sterile conditions at 37°C in a humidified incubator with 5% CO_2_ unless otherwise specified.

#### Quantification of p62-positive aggregates in iPSC-derived neural stem cells

Cryopreserved NSCs were thawed at passage 2 and expanded to confluence, then passaged and plated on black-walled, clear-bottom 96-well Geltrex-coated plates (Costar, 3904) in Neural Expansion Medium (NEM; 50% Neurobasal medium, 50% Advanced DMEM/F-12, 2% Neural Induction Supplement, and 1% penicillin–streptomycin) containing ROCK inhibitor Y-27632 (1:2,000; Selleck Chemicals, S1049) on day 0. Media were exchanged with fresh NEM on day 1and the cells were treated overnight (∼18 h) with NEM containing 0, 1, 10, or 100 µM chloroquine (Cell Signaling Technology, 14774). Each cell line and treatment condition were tested in quadruplicate. For inter-plate normalization, two cell lines were plated in triplicate on each plate and treated with 100 µM chloroquine. All experiments were performed using NSCs at passages 3–5.

Following treatment, cells were fixed in 4% paraformaldehyde (Electron Microscopy Sciences, 15710) in 1× PBS for 20 min, washed three times with PBS, and blocked/permeabilized for 1 h in PBS containing 0.5% Triton X-100 and 5% bovine serum albumin (BSA; Sigma-Aldrich, A7906). Cells were incubated overnight at 4°C with mouse monoclonal anti-SQSTM1/p62 (Abcam, ab56416; 1:500) diluted in PBS containing 2.5% BSA and 0.25% Triton X-100. The next day, cells were washed twice with PBS and incubated for 1 h with donkey anti-mouse IgG Alexa Fluor™ Plus 488 secondary antibody (Invitrogen, A32766; 1:1,000) in PBS with 2.5% BSA and 0.1% Triton X-100. Nuclei were stained with NucBlue™ (ThermoFisher Scientific, R37606; 2 drops/mL PBS) for 30 min and washed three times with PBS.

Image acquisition and analysis were performed using the ImageXpress Confocal HT.ai (Molecular Devices) with laser-based autofocus at 20× magnification. Nine sites were imaged per well using MetaXpress High-Content Image Acquisition and Analysis software (v6.7.2.290). Images were analyzed using the granularity application module, where nuclei were segmented by NucBlue fluorescence and p62-positive puncta were identified as FITC fluorescence exceeding 500 gray levels above local background and with widths between 1 and 10 µm. The total granule area per cell was averaged across nine sites per well. For each experimental batch, the mean granule area per cell from the two cell lines plated on each plate was used for inter-plate normalization.

Statistical analyses were performed in GraphPad Prism v10.5.0 (GraphPad Software, San Diego, CA). Two-way analysis of variance (ANOVA) was used to test for main effects of chloroquine treatment and genotype, followed by Tukey’s post hoc test for multiple comparisons.

#### Tau seeding assays in wild-type isoform-specific and P301S tau biosensor cell lines

For tau seeding assays assessing isoform-specific aggregation, we employed HEK293T-based 4R-only and mixed 3R/4R wild-type (WT) tau biosensor cell lines engineered to report intracellular tau aggregation via FRET^57^. The 4R-only biosensor line constitutively expresses the human WT tau repeat domain (RD; residues 246–408) containing four microtubule-binding repeats, C-terminally fused to monomeric mCerulean3 and monomeric mRuby3 (RD(4Rext)-Cer/Rub). The mixed-isoform biosensor line expresses the 3R WT tau RD fused to mCerulean3 and the 4R WT tau RD fused to mRuby3 (RD(3R/4Rext)-Cer/Rub).

As described previously^57^, biosensor cells were plated in 96-well plates at a density of 20,000 cells per well and maintained in Dulbecco’s Modified Eagle Medium (DMEM) supplemented with 10% fetal bovine serum and 1% penicillin-streptomycin. XDP and control brain lysates were sonicated and applied to cells at a final concentration of 7.5 µg total protein per well. For transfection, lysates were mixed with 0.50 µL Lipofectamine™ 2000 and 9.50 µL Opti-MEM™ and incubated at room temperature for 30 min prior to addition to the cells. Each condition was performed in triplicate wells. Cells were incubated for 72 h to permit seeded aggregation, after which they were harvested and analyzed by flow cytometry to quantify FRET-positive populations as a measure of tau seeding activity.

In parallel, we employed V2L tau biosensor cells (NK13) expressing the human tau repeat domain harboring the disease-associated P301S mutation, fused to mCerulean3 or mClover3 (tau RD-Cer/Clo), enabling detection of intracellular tau aggregation via FRET^55^. V2L biosensor cells were plated in 96-well plates at a density of 20,000 cells per well and maintained in DMEM supplemented with 10% fetal bovine serum and 1% penicillin–streptomycin. Brain lysates were sonicated for 5 min using 30 s on/off cycles at an amplitude of 65 with a Qsonica sonicator and 10 μg of total protein from the soluble fraction was added per well, whereas 3 μL of the sonicated insoluble pellet suspension was added per well. For transfection, lysates were mixed with 0.5 µL Lipofectamine™ 2000 and 9.5 µL Opti-MEM™ and incubated at room temperature for 20 min prior to addition to the cells. Each condition was performed in triplicate wells. Cells were incubated for 48 h, after which they were harvested and analyzed by flow cytometry to quantify FRET-positive populations.

### Molecular analyses

#### RNA extraction, sequencing, and preprocessing

Total RNA was extracted from frozen brain tissue by bead-based homogenization in TRIzol™ reagent (ThermoFisher Scientific) followed by phase separation and ethanol precipitation. RNA quantity and integrity were assessed using an Agilent Bioanalyzer, and only samples with RNA integrity number (RIN) ≥ 6 were used for library preparation. Poly(A)-enriched, stranded mRNA-seq libraries were constructed using the Illumina TruSeq Stranded mRNA kit and sequenced on an Illumina NovaSeq X platform, generating paired-end 150-bp reads (minimum 25 million, maximum 107 million reads per sample).

Raw FASTQ files were assessed with FastQC and RNA-SeqQC^105^, and adapter sequences were trimmed using Trimmomatic v0.36. Reads were aligned to the human reference genome (GRCh38) using STAR v2.7.10133 with the following parameters: --outFilterMultimapNmax 1 --outFilterMismatchNoverLmax 0.05 --alignEndsType EndToEnd. Gene-level counts were obtained directly from STAR. Intron retention in TAF1 intron 32 was quantified using IRFinder^106^. Quality-control metrics for all samples are provided in **Table S1**.

#### Differential expression and enrichment analyses

Differential gene-expression analyses were performed in R using DESeq2 v1.46.0 for each brain region separately (striatum: 28 XDP, 10 controls; cerebellum: 37 XDP, 13 controls; prefrontal cortex: 36 XDP, 11 controls; occipital cortex: 34 XDP, 13 controls; all males; **Table S1**). Genes with counts > 1 cpm across samples were retained, resulting in 13,878 (striatum), 13,943 (cerebellum), 13,687 (prefrontal cortex), and 13,495 (occipital cortex) genes analyzed. Normalization used the median-of-ratios method^128^, and surrogate variables were estimated via SVAseq v3.54.0108 to adjust for technical variation. Differentially expressed genes (DEGs) were defined at FDR-adjusted p<0.01 and |log_2_ fold-change|>0.58. Gene symbols were annotated using AnnotationDB v1.6.8. Full DESeq2 outputs are provided in **Table S3**.

To identify overlaps among regional DEG lists, direction-specific hypergeometric tests were performed using genes with adj-*p*<0.01. Backgrounds were defined by the set of expressed genes per region. Marker genes for striatal medium spiny neurons (related to **Figure 4B**) were obtained from the literature^60^. For analyses of expression changes associated with (CCCTCT)_n_ repeat length, DEGs identified in the striatum were regressed against repeat length among XDP samples using a linear model (*p*<0.05; Table S4).

Functional enrichment of gene sets—including DEGs, overlapping DEGs, and repeat-length–associated DEGs—was performed using gprofiler2 v0.2.3, querying “GO:BP,” “GO:CC,” “GO:MF,” “KEGG,” “REAC,” and “WP” ontologies. Backgrounds were limited to the expressed genes per region, and enrichments with FDR-adjusted *p*<0.05 were retained. Curated enrichments represented in **Figure 4A** are summarized in **Table S5**. Literature-based enrichment analyses used Fisher’s exact tests. Overlaps with HD medium spiny neuron signatures were computed using C-phase gene statistics from Handsaker et al.^60^, and co-expression modules from Alzheimer’s-disease prefrontal cortex^68^ were compared to those from XDP prefrontal cortex networks.

#### Cell-type deconvolution and coexpression network analysis

Cell-type deconvolution was performed using CIBERSORTx^129^. Bulk RNA-seq profiles from striatum and prefrontal cortex were analyzed in “relative” mode using reference transcriptomes from a published benchmarking dataset for brain tissue deconvolution^130^. Cell-type fractions were compared between XDP and controls using independent *t*-tests.

Weighted gene coexpression network analysis (WGCNA v1.3)^131^ was conducted on normalized striatal and prefrontal cortex expression matrices. Log-transformed counts were used to generate signed networks based on adjacency and topological overlap matrices (soft-threshold = 20). Modules with ≥50 genes were identified. Genes with module-membership *p*>0.05 were reassigned to the unclassified module (M0; 1,689 genes for prefrontal cortex, 1,707 genes for striatum). Module eigengenes were correlated with traits (levels of intron retention/IR, staining for Ubiquitin, p62, p-tau181, p-tau202/205, p-tau231, p-tauT17, total tau (Tau5) and misfolded tau (T22), and GFAP (striatum only), **Table S7**), and enrichment of DEGs within modules was tested via hypergeometric tests. Modules showing eigengene-trait correlations FDR<0.1 and DEG enrichment FDR<0.01 were included in **Figure 4C-D**.

Genotyping for MAPT H1/H2 haplotypes was performed using the TaqMan™ SNP Genotyping Assay for rs9468, which discriminates the H1 and H2 haplotypes through allele-specific fluorescent probes (VIC dye for the “T” allele [H1], FAM dye for the “C” allele [H2]). Each 20 µL reaction contained 10 µL TaqMan™ Fast Advanced Master Mix (ThermoFisher Scientific, 4444557), 1.0 µL TaqMan™ SNP Genotyping Assay for rs9468 (ThermoFisher Scientific, 4351379), 7.0 µL nuclease-free water, and 2 µL genomic DNA (10-20 ng).

Quantitative PCR was performed on a StepOne™ Plus Real-Time PCR System under standard cycling conditions recommended by the manufacturer: one cycle at 25 °C for 30 s, one cycle at 95 °C for 20 s, followed by 40 cycles of 95 °C for 3 s and 60 °C for 20 s, and a final hold at 25 °C for 20 s. Allelic discrimination was conducted using Applied Biosystems QuantStudio™ software, and haplotype assignments were confirmed by manual inspection of fluorescence clustering.

### Plasma profiling

#### Immunoassay measurements

Quantification of plasma biomarkers was performed at the CTRU Biomarker Core at MG using Mesoscale Discovery (MSD) electrochemiluminescence immunoassays. Individual assays were used for phosphorylated tau181 (p-tau181; K151AGMS), fractalkine (K151VCK-2), and YKL-40 (K151VLK-2), and a multiplex 3-plex assay (K15639S) was used for glial fibrillary acidic protein (GFAP), total tau (tTau), and neurofilament light chain (NfL). Assays for 156 individuals (120 XDP, 36 controls) were performed. Each sample was plated in duplicate and randomized across plates; plate medians were used for normalization. The median coefficient of variation (CV) across duplicates was <6% for all markers (NfL, 5.75%; GFAP, 4.2%; tTau, 2.6%; p-tau181, 6.0%; fractalkine, 2.9%; YKL-40, 2.5%). Samples with CV>20% were excluded from analyses.

#### Statistical analyses of plasma biomarkers

To account for differences in age at collection of plasma samples, the cohort was restricted to the subset that were collected between ages 30-60 (n=119 XDP, 29 Controls), then genotype differences per biomarker were tested by applying linear regression to the log-transformed biomarker values (log(biomarker) ∼ age + genotype). Log-transformation was used since the biomarker values are positive-valued and skewed to the right. To assess sensitivity of these results to the linear regression assumptions, we also applied rank-based ANCOVA (using Rfit in R) with the same model (log(biomarker) ∼ age + genotype)) as a nonparametric test of differences between cases and controls, adjusting for age (**Figure S10**). Age-adjusted residuals from rank-based ANCOVA were also applied to construct receiver operator characteristic (ROC) curves. ROC analyses were performed in R (v4.1.3) using the pROC package (v1.19.0.1).

#### Statistical analyses

All statistical analyses (when not stated) were performed in GraphPad Prism v10.5.0 (GraphPad Software, San Diego, CA).

---

**Figure S1.**
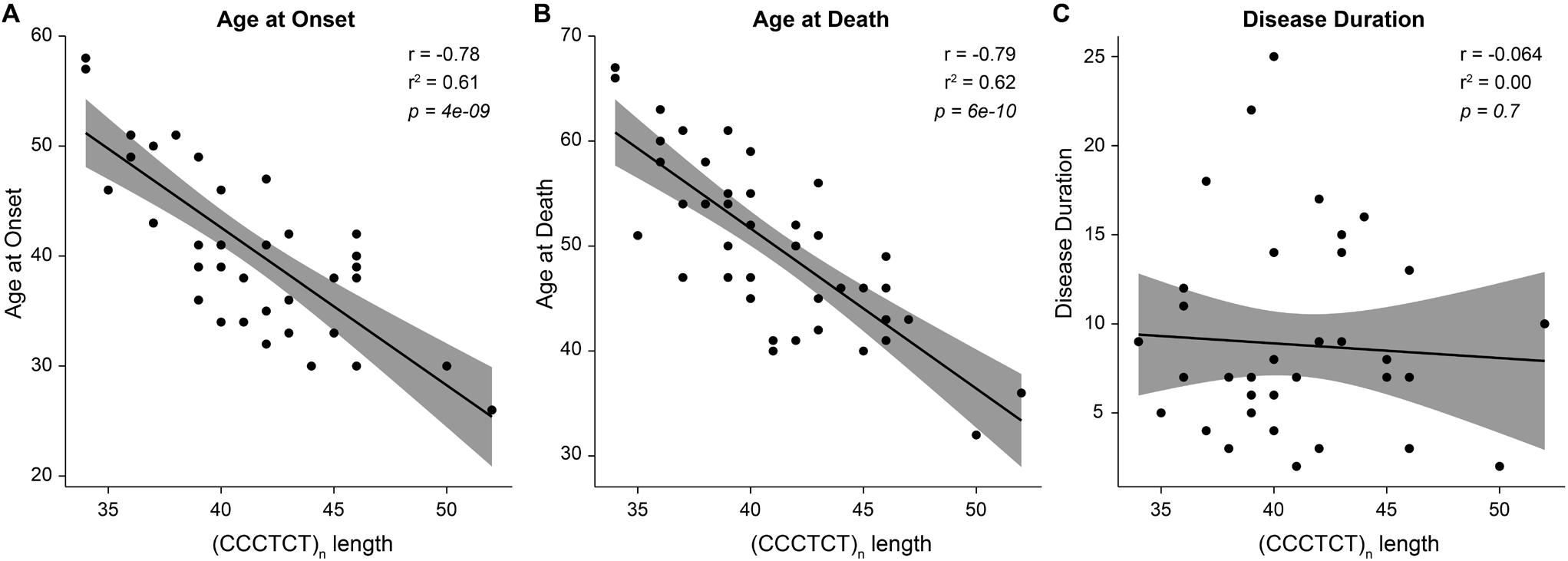
Correlations between XDP-specific (CCCTCT)_n_ repeat length, clinical features, and cohort characteristics. **A-B**, (CCCTCT)_n_ repeat length shows strong inverse correlations with age at onset and age at death in patients with XDP. **C**, No significant correlation is observed between repeat length and disease duration. Repeat length ranges from 34-52 and age at onset from 26-58 years. Post-mortem interval and RNA integrity values do not differ between XDP and control brains (see **Table S1**).

**Figure S2.**
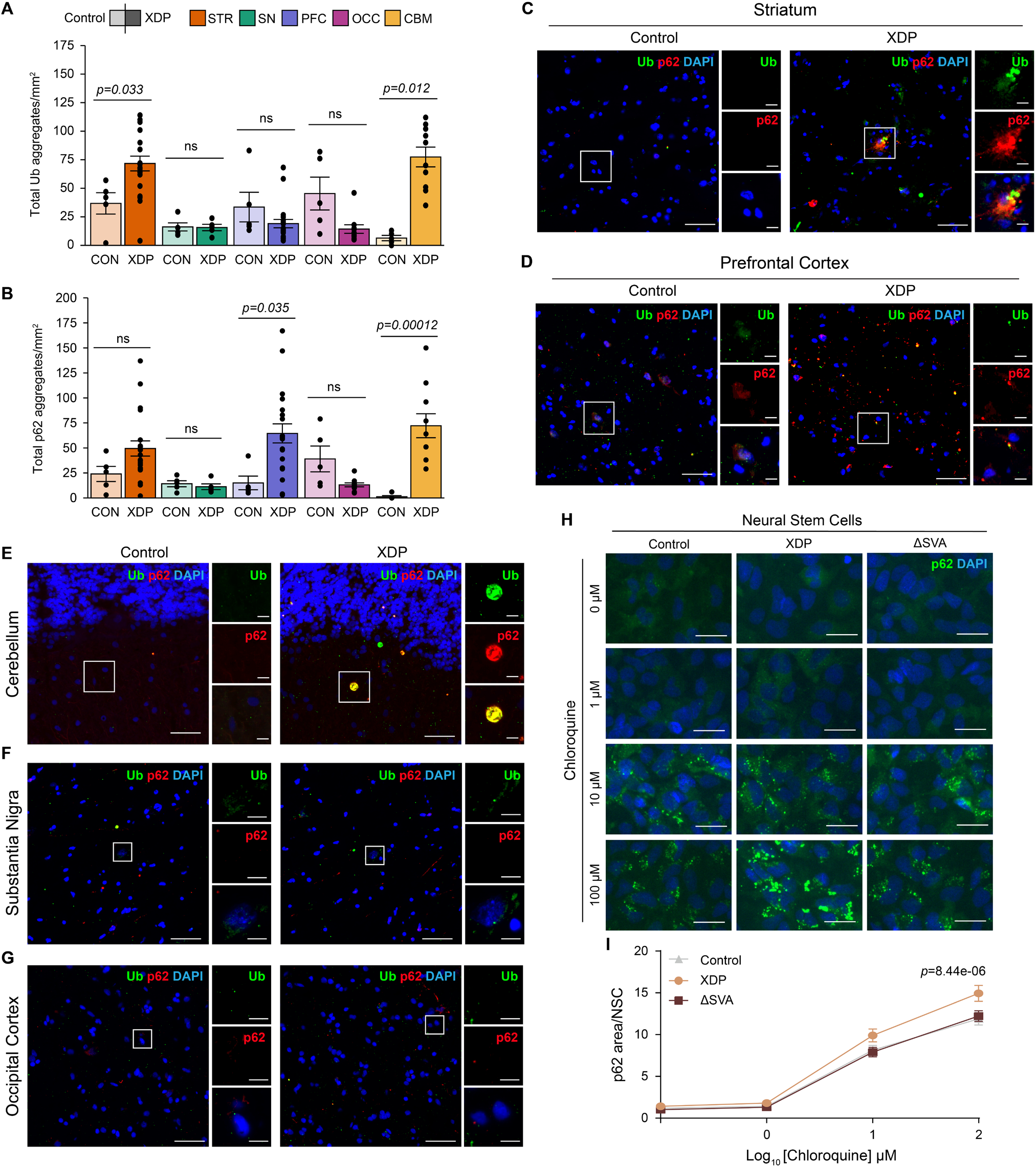
Region-specific accumulation of ubiquitin- and p62-positive aggregates and increased p62 levels upon autophagic inhibition in XDP. **A-B**, Quantification of total ubiquitin (Ub) (**A**) and p62 (**B**) aggregates per mm^2^ across brain regions shows increased Ub-positive aggregates in the striatum (STR) and cerebellum (CBM), and increased p62-positive aggregates in the prefrontal cortex (PFC) and CBM in X-linked dystonia-parkinsonism (XDP) compared to control. No significant change was observed in substantia nigra (SN) or occipital cortex (OCC). **C**, Representative confocal images of control and XDP STR stained for Ub and p62. Insets show higher-magnification views of boxed regions. **D**, Representative images of control and XDP PFC stained for Ub and p62. **E**, Representative images of control and XDP CBM stained for Ub and p62, showing accumulation of inclusions in the molecular layer. **F**, Representative images of control and XDP substantia nigra (SN) stained for Ub and p62, showing no significant changes. **G**, Representative images of control and XDP occipital cortex (OCC) stained for Ub and p62, showing no significant differences. **H**, Representative images of neural stem cells (NSCs) derived from control (n=5), XDP (n=6), and isogenic SVA-deleted (ΔSVA) XDP lines (n=6) treated with increasing concentrations of chloroquine (0-100 μM) to inhibit autophagy, showing dose-dependent accumulation of p62-positive puncta. **I**, Quantification of p62-positive area per cell shows that p62 levels are elevated in XDP NSCs relative to control and ΔSVA lines upon autophagic inhibition. Data are mean ± s.e.m.; each point represents one case or independent cell line. Statistical significance is indicated in panels. Scale bars, 50 μm (tissue images) and 10 μm (insets and NSCs).

**Figure S3.**
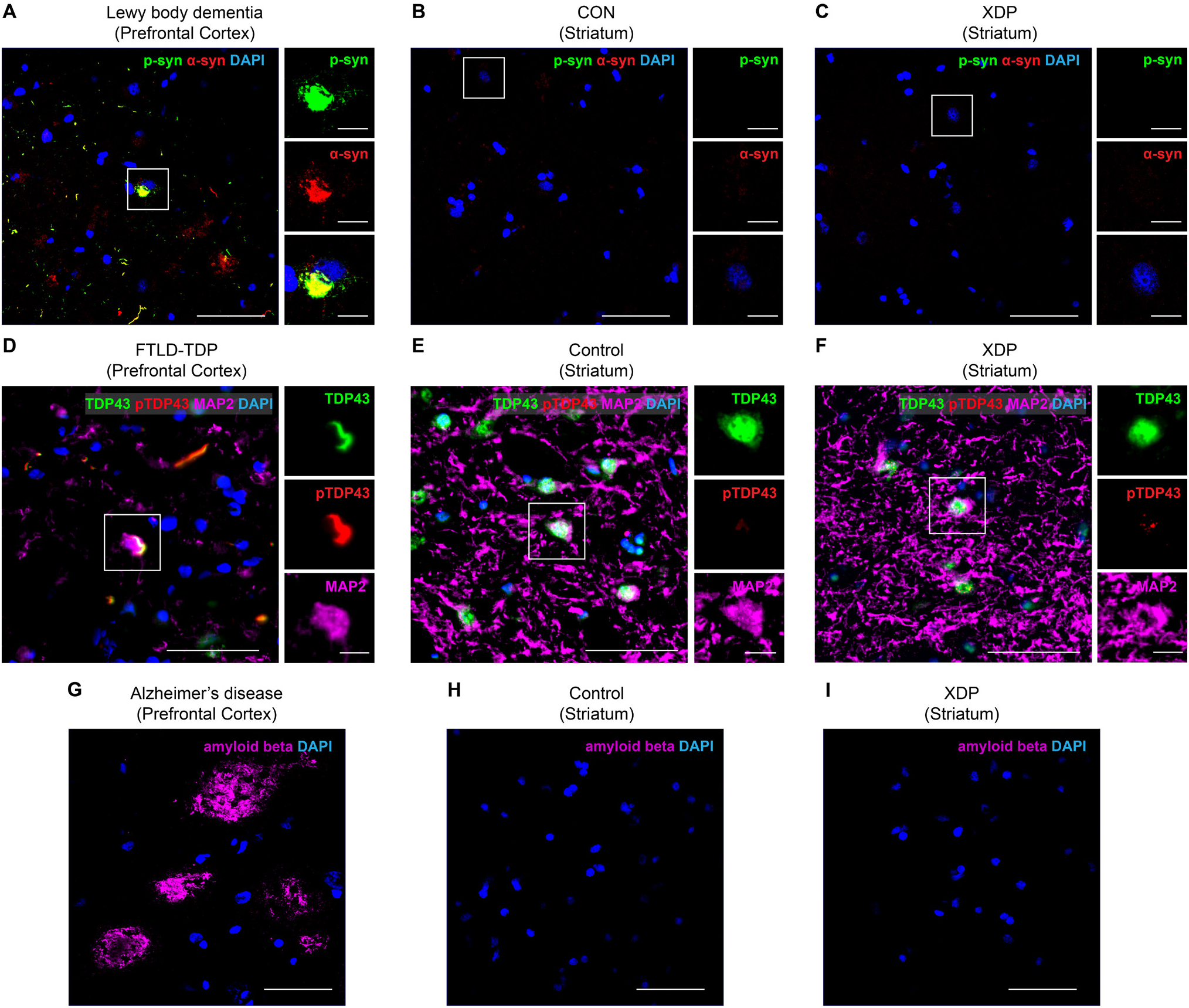
Absence of α-synuclein, TDP-43, and amyloid-β pathology in XDP brains. **A-C**, Representative confocal images stained for α-synuclein (α-syn) and phosphorylated α-synuclein (p-syn; Ser129) in dementia with Lewy bodies (DLB) prefrontal cortex (**A**, positive control), control striatum (**B**), and X-linked dystonia-parkinsonism striatum (**C**). Robust Lewy body-like inclusions are observed in DLB, whereas no abnormal α-synuclein deposition is detected in control or XDP brains. Insets show higher-magnification views of boxed regions. **D-F**, Representative images stained for total TDP-43, phosphorylated TDP-43 (pTDP-43), and MAP2 in prefrontal cortex of frontotemporal lobar degeneration with TDP-43 pathology (FTLD-TDP) (**D**, positive control), control striatum (**E**), and XDP striatum (**F**). FTLD-TDP shows nuclear clearance and cytoplasmic inclusions of TDP-43, whereas control and XDP brains show preserved nuclear localization without pathological inclusions. Insets show higher-magnification views. **G-I**, Representative images of amyloid-β immunostaining in prefrontal cortex of Alzheimer’s disease (**G**, positive control), control striatum (**H**), and XDP Sstriatum (**I**). Amyloid plaques are evident in Alzheimer’s disease, whereas no amyloid-β deposition is detected in control or XDP brains. These findings indicate the absence of co-pathologies associated with α-synuclein, TDP-43, and amyloid-β aggregation in XDP.

**Figure S4.**
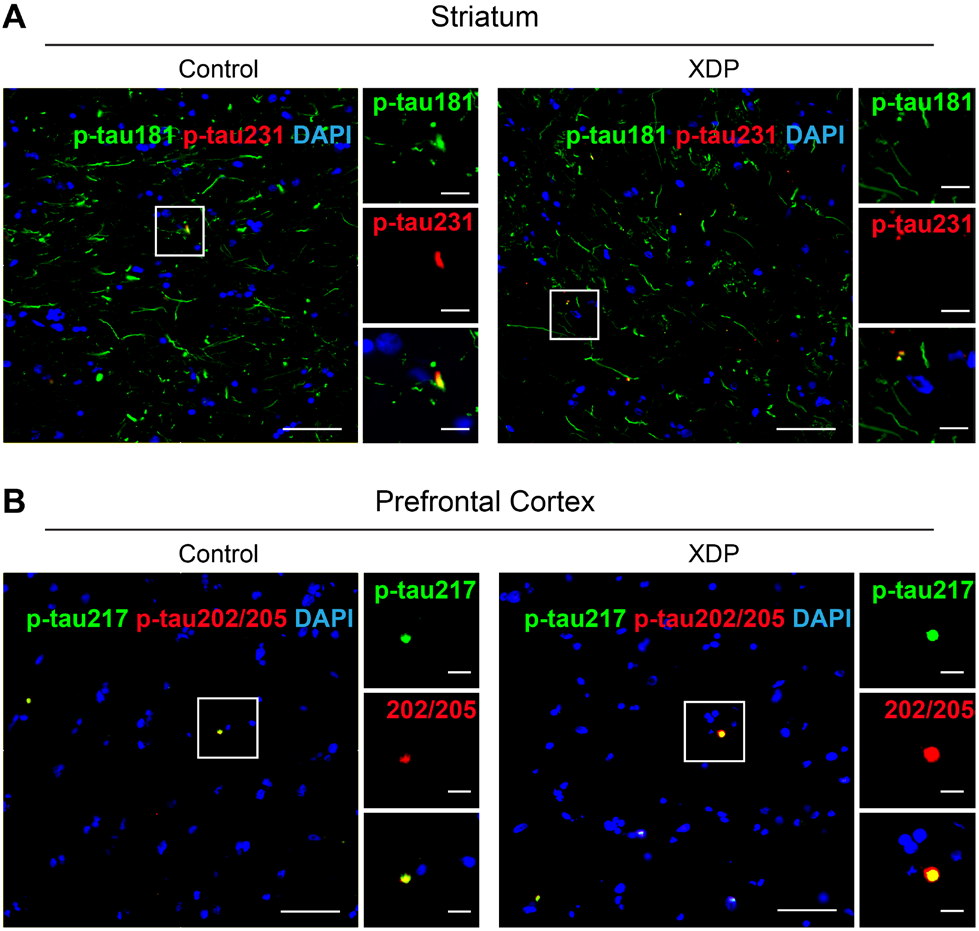
No detectable changes in specific phospho-tau markers in the striatum and prefrontal cortex in XDP. **A**, Representative confocal images of control and XDP striatum stained for p-tau181 and p-tau231, showing no detectable differences between groups. Insets show higher-magnification views of boxed regions. **B**, Representative images of control and XDP prefrontal cortex stained for p-tau217 and p-tau202/205 (AT8), showing no detectable differences between groups. Insets show higher-magnification views. Scale bars, 50 μm (overview images) and 10 μm (insets).

**Figure S5.**
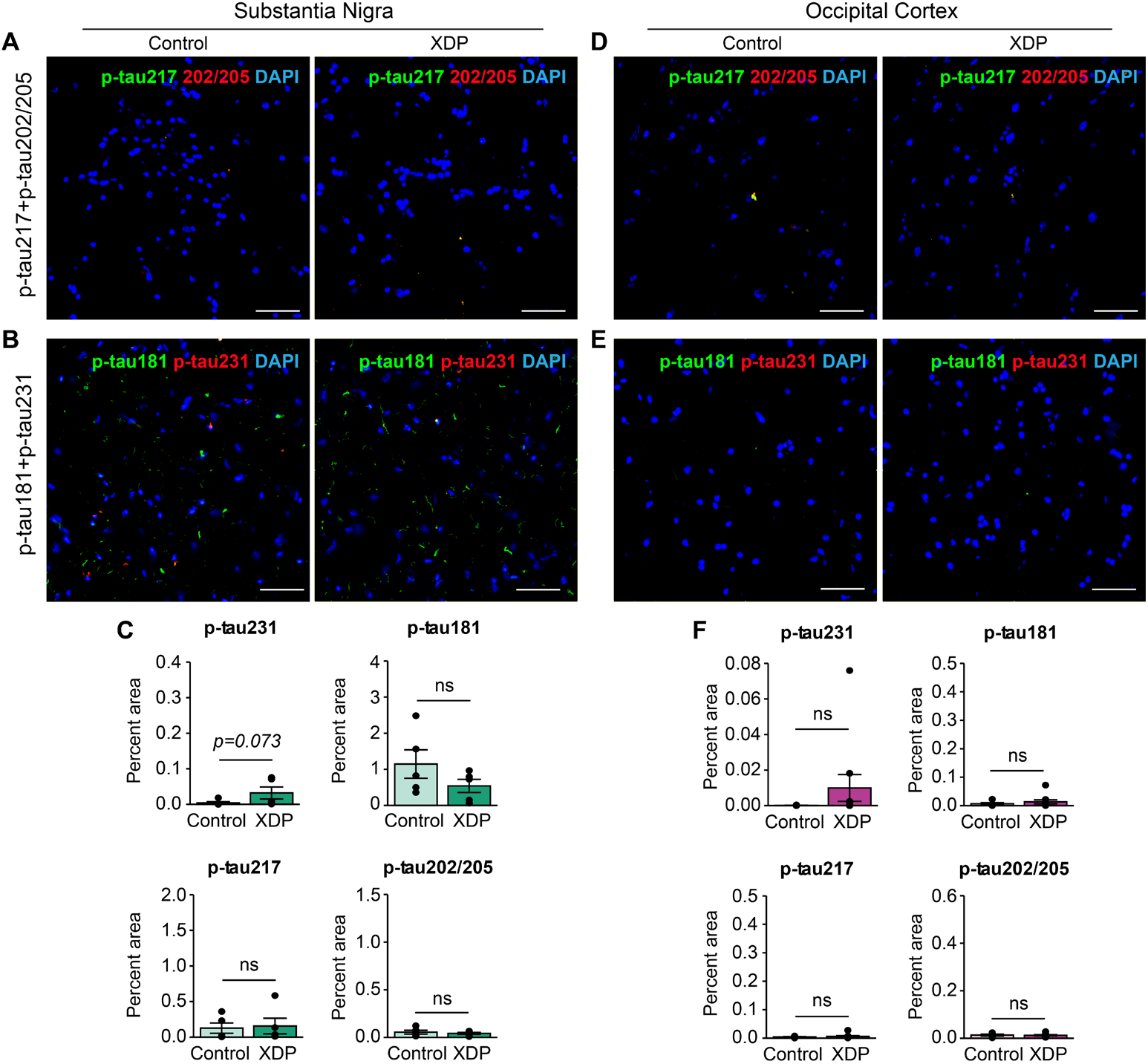
No detectable changes in phospho-tau markers in the substantia nigra and occipital cortex in XDP. **A-B**, Representative confocal images of control and XDP substantia nigra stained for p-tau217 and p-tau202/205 (AT8) (**A**) and p-tau181 and p-tau231 (**B**), showing no detectable differences between groups. **C**, Quantification of phospho-tau burden in the substantia nigra (p-tau231, p-tau181, p-tau217, and p-tau202/205) shows no significant differences between control and XDP. **D-E**, Representative images of control and XDP occipital cortex stained for p-tau217 and p-tau202/205 (AT8) (**D**) and p-tau181 and p-tau231 (**E**), showing no detectable differences between groups. **F**, Quantification of phospho-tau burden in the occipital cortex confirms no significant differences across all epitopes examined. Insets show higher-magnification views of boxed regions. Scale bars, 50 μm (overview images) and 10 μm (insets).

**Figure S6.**
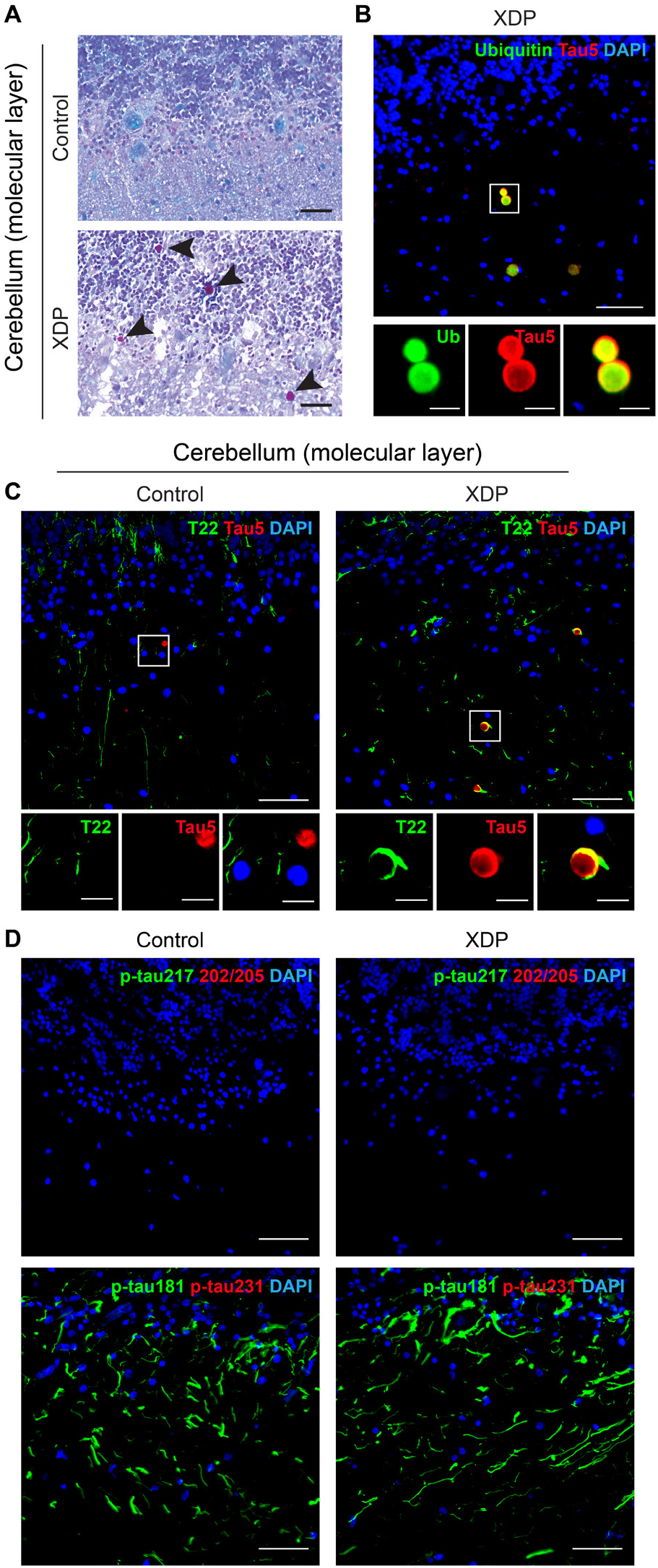
Tau-immunoreactive corpora amylacea accumulate in the cerebellum of patients with XDP. **A**, Periodic acid-Schiff (PAS) staining of control and XDP cerebellum highlights the polysaccharide-rich core of corpora amylacea (arrowheads), which are prominent in XDP. **B**, Confocal image of an XDP cerebellum stained for ubiquitin (Ub) and total tau (Tau5) shows that total tau is present within ubiquitin-positive corpora amylacea. Insets show higher-magnification views of boxed regions, demonstrating co-localization. **C**, Representative confocal images of control and XDP cerebellum stained for oligomeric tau (T22) and total tau (Tau5). Tau5-positive spherical structures consistent with corpora amylacea are observed, with minimal T22 signal occasionally staining the periphery. Insets show higher-magnification views. **D**, Representative images of control and XDP cerebellum stained for p-tau217 and p-tau202/205 (AT8) (top) and p-tau181 and p-tau231 (bottom), showing absence of phospho-tau immunoreactivity within corpora amylacea. Data are representative of independent cases. Scale bars, 50 μm (overview images) and 10 μm (insets).

**Figure S7.**
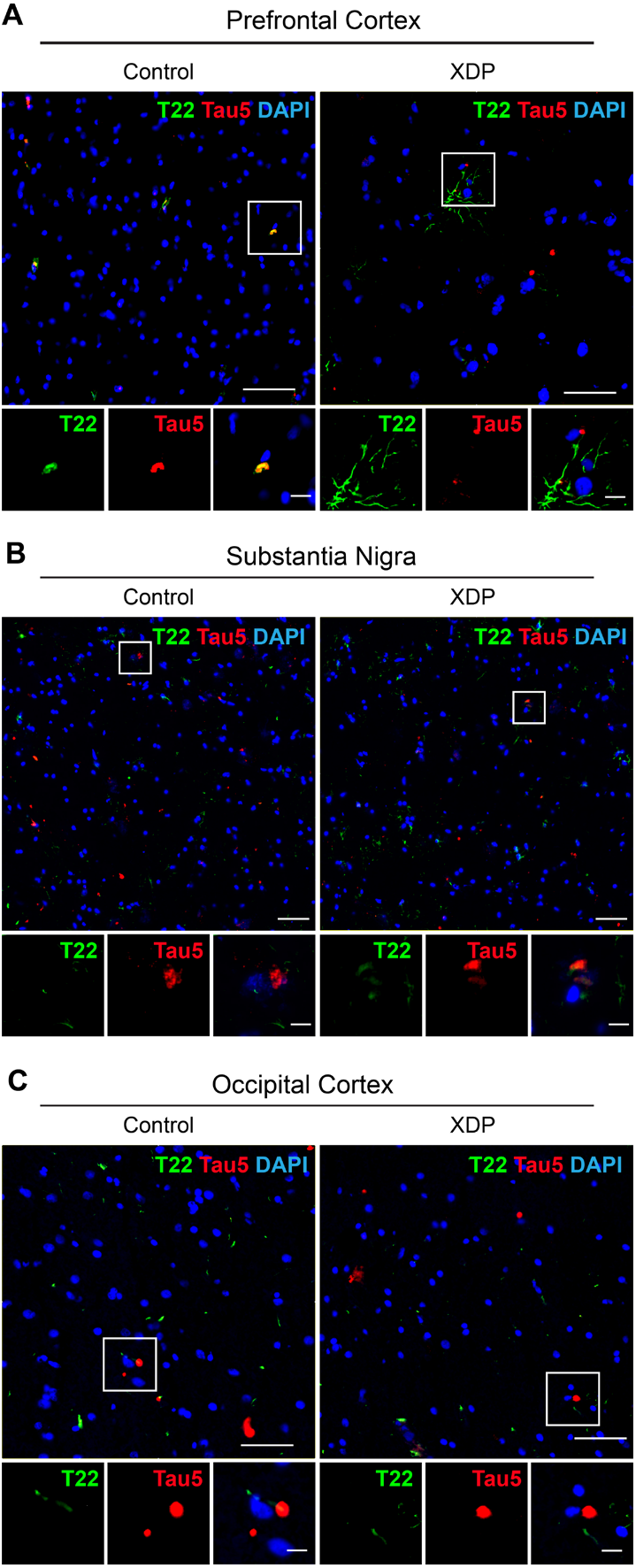
Oligomeric tau immunoreactivity in extra-striatal brain regions of patients with XDP. **A-C**, Representative confocal images of the prefrontal cortex (A), substantia nigra (B), and occipital cortex (C) from control and X-linked dystonia-parkinsonism (XDP) cases stained for oligomeric tau (T22; green), total tau (Tau5; red), and nuclei (DAPI; blue). Boxed regions are shown at higher magnification below each panel. Occasional T22-positive structures were observed in both control and XDP brains across these regions. Data are representative of independent cases. Scale bars, 50 μm (overview images) and 10 μm (insets).

**Figure S8.**
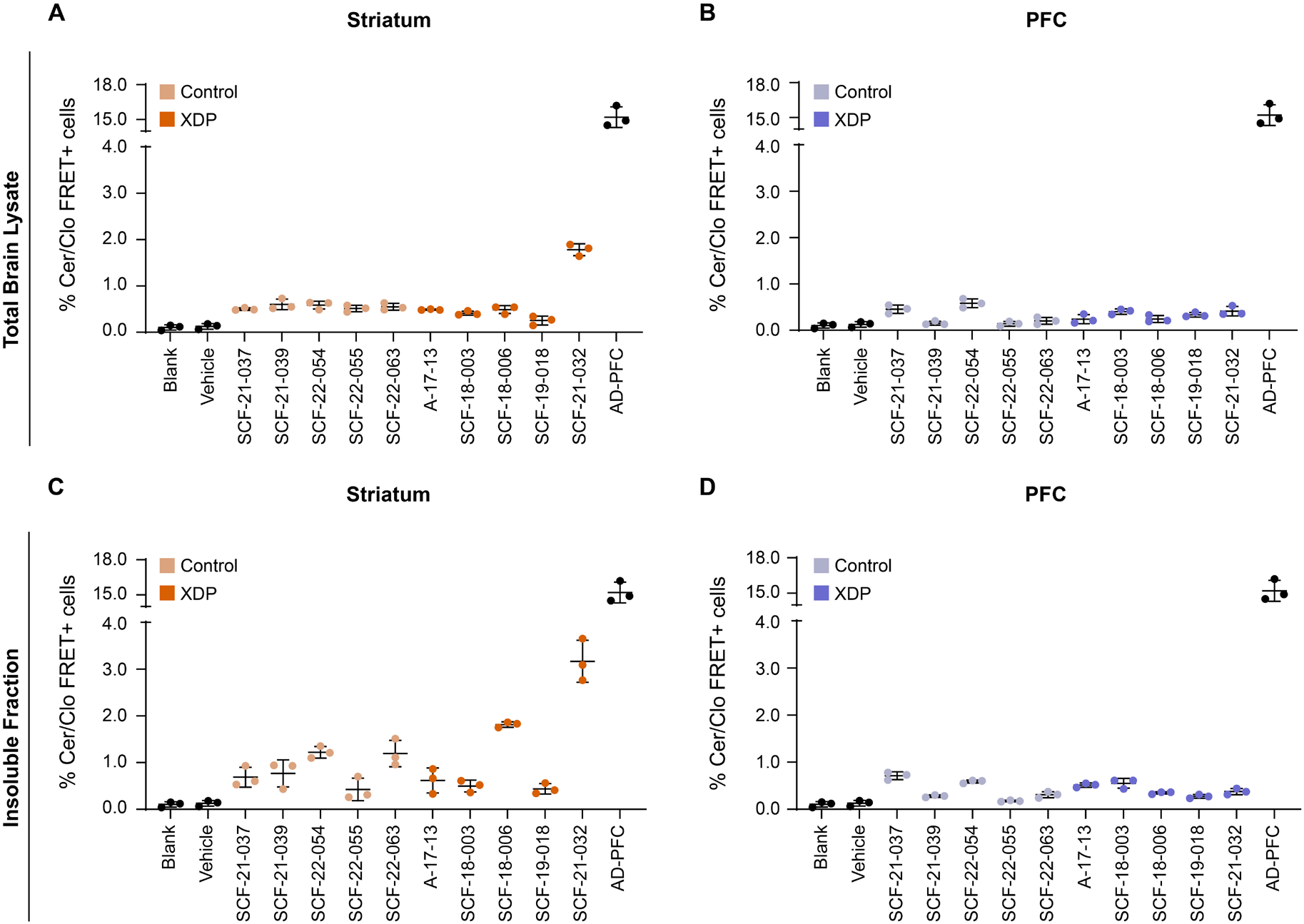
Individual case-level tau seeding activity in soluble and insoluble fractions assessed using the ultrasensitive v2L biosensor. **A-D**, Tau seeding activity of individual control and X-linked dystonia-parkinsonism (XDP) cases following transfection of total striatal (**A**) and prefrontal cortex (PFC) (**B**) lysates, and insoluble striatal (**C**) and prefrontal cortex (**D**) fractions into HEK293T v2L tau biosensor cells. The vehicle-only control consisted of Lipofectamine transfection in the absence of brain lysate. No detectable increase in tau seeding activity was observed in TBS-soluble or sarkosyl-insoluble PFC extracts from either control or XDP cases (**B, D**). Alzheimer’s disease (AD) PFC lysates served as positive controls. Data are presented as mean ± s.e.m., with each point representing an independent technical replicate.

**Figure S9.**
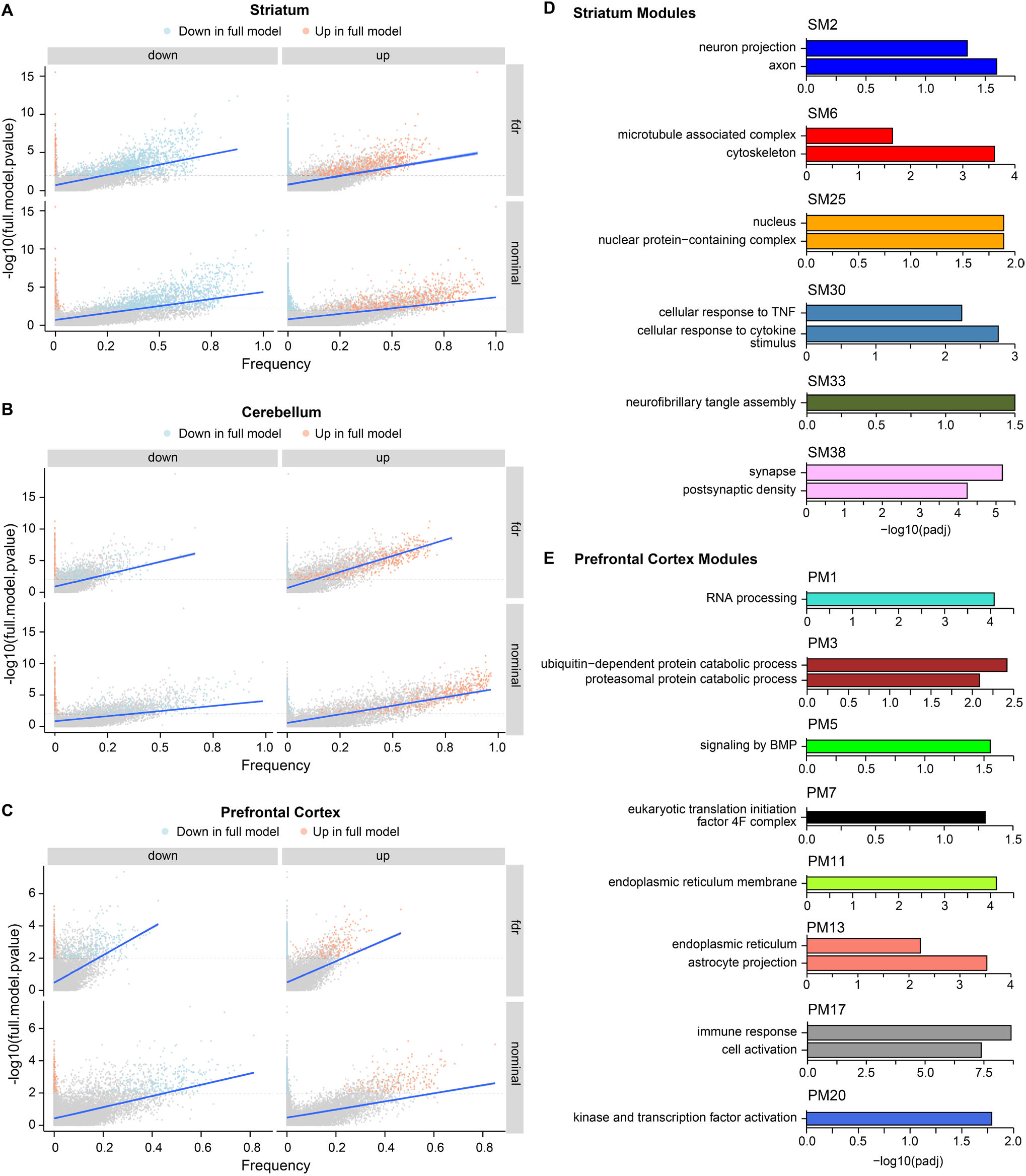
Robust differential expression and co-expression module enrichments across brain regions in XDP. **A-C**, Bootstrapped differential gene expression analyses. Differentially expressed genes (DEGs, adj-*p*<0.01, |log_2_FC| > 0.58) identified in the full dataset are consistently recovered (>20% of bootstraps) across 50,000 resampled Wald tests using six XDP and six control profiles each (per brain region), indicating robust full model analyses despite imbalance of cases and controls. **D-E**, Enrichments in co-expression modules. Functional enrichments in selected co-expression modules from the prefrontal cortex and striatum recapitulate dysregulated processes observed from the differential expression analyses and exemplify tau-related signatures also found in the literature.

**Figure S10.**
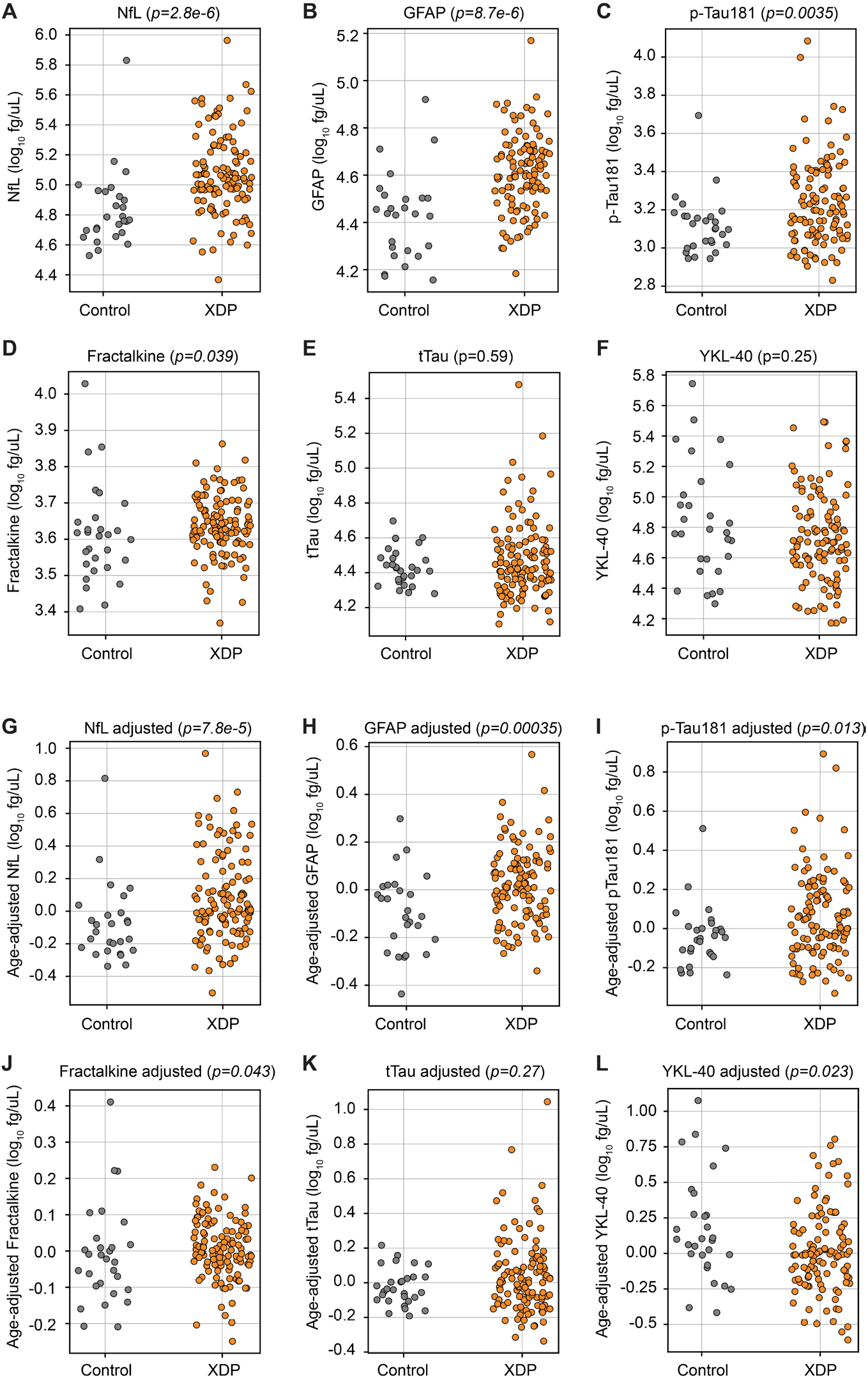
Plasma biomarker differences between XDP and controls before and after adjustment for age. **A-F**, Plasma concentrations of neurofilament light chain (NfL) (**A**), glial fibrillary acidic protein (GFAP) (**B**), phosphorylated tau-181 (p-tau181) (**C**), fractalkine (**D**), total tau (tTau) (**E**), and YKL-40 (**F**) in control (n=29) and X-linked dystonia-parkinsonism (XDP; n=119) participants from the age-restricted cohort (30-60 years at plasma collection). Biomarker values were log-transformed prior to statistical testing to account for right-skewed distributions. **G-L**, Age-adjusted biomarker values derived from rank-based ANCOVA using the model log(biomarker) ∼ age + genotype. NfL (**G**), GFAP (**H**), p-tau181 (**I**), fractalkine (**J**), and YKL-40 (**L**) remained significantly altered in XDP after adjusting for age, whereas tTau (**K**) did not differ between groups. Corresponding *p* values from rank-based ANCOVA are indicated in each panel. Age-adjusted residuals from these analyses were subsequently used to construct receiver operating characteristic (ROC) curves in **Figure 7**. Each point represents one participant.

## Notes

### Author Declarations

Ethics committee/Institutional Review Board of Massachusetts General Brigham (Boston, MA, USA) gave ethical approval for this work. Institutional Review Board of Makati Medical Center (Makati, National Capital Region, Philippines) gave ethical approval for this work. Institutional Review Board of West Visayas State University (Iloilo City, Iloilo, Philippines) gave ethical approval for this work. Institutional Review Board of Jose R. Reyes Memorial Medical Center (Manila, Philippines) gave ethical approval for this work.

